# Integrative kidney multi-omics traces genetic drivers of chronic kidney disease to targets within the renal epigenome, transcriptome and proteome

**DOI:** 10.64898/2026.01.15.26344229

**Authors:** Amber Emmett, Xiaoguang Xu, Xiao Jiang, Shadi Hames-Fathi, David Scannali, James Eales, Ewa Miller-Kasprzak, Polly Downton, Antony Adamson, Yong Sun, Abigail C. Lay, David Talavera, Matthew Denniff, John Dormer, Grzegorz Rempega, Robert Król, Sebastien Rubin, Joanna Zywiec, Wojciech Wystrychowski, Pawel Bogdanski, Cristian Pattaro, Nilesh J. Samani, Bernard Keavney, Tomasz J. Guzik, Rachel Lennon, Andrew P. Morris, Fadi J. Charchar, Maciej Tomaszewski

## Abstract

Chronic Kidney Disease (CKD) is a complex polygenic disease. We performed genome-wide association meta-analyses of four CKD-defining traits in up to 890,000 individuals and identified 508 loci associated with at least one trait, including 237 multi-trait CKD loci. Colocalization with kidney mRNAs, proteins and methylation patterns prioritized 651 CKD kidney genes (including 330 novel candidates) at 320 CKD-defining trait loci. We discovered over-representation of CKD kidney genes within multi-trait CKD loci. CKD kidney genes which colocalized with multiple CKD-defining traits exhibited greater relevance to kidney biology, health and disease. We found evidence for genetic regulation of developmental DNA methylation patterns that determine kidney health later in life. Finally, through analysis of Isolated Hyperchlorhidrosis - a rare genetic syndrome associated with *Carbonic Anhydrase 12* (a novel CKD kidney gene) - we uncovered new metabolic consequences of genetic *CA12* loss (hyperuricemia, reduced kidney function) and illuminated adverse effects of CA12-inhibitors (acetazolamide).

## Introduction

Chronic kidney disease (CKD) affects ∼10% of adults and is a complex, heritable and polygenic trait^1^. CKD is an increasing burden to healthcare and mortality across the globe and is predicted to be the fifth most common cause of years of life lost by 2040^2^. Genome-wide association studies (GWAS) have uncovered common genetic variants associated with CKD^3–7^ through analysis of its defining quantitative traits: estimated glomerular filtration rate by creatinine (eGFRcr) or cystatin C (eGFRcys), blood urea nitrogen (BUN), and urinary albumin creatinine ratio (UACR). Leveraging multi-omics data, post-GWAS gene prioritization studies have elucidated the molecular underpinnings of CKD; translating GWAS signals into genes, molecules and pathways^5,8–14^. Despite a spectacular success of these studies in enhancing our understanding of the molecular drivers of CKD, a few outstanding challenges remain to be addressed.

Firstly, most of the previously identified drivers of CKD come primarily from GWAS of eGFRcr as the CKD-defining trait. Indeed, a majority of the previous studies relied on either eGFRcr as the only biochemical marker of kidney function^3^ or prioritized eGFRcr signals coinciding with weak associations with eGFRcys or BUN (typically *P* < 0.05 with the lead GWAS variant)^5,11,12^. These eGFRcr-centric approaches may have missed associations with non-creatinine-based kidney function measurements and/or enhanced the signals driven primarily by creatinine metabolism. Instead, post-GWAS studies that leverage multiple CKD-defining traits^8,13^ are better designed to capture the endophenotype of kidney disease susceptibility, which is proxied by the convergence of multiple measurements of kidney function and damage, mirroring the diagnostic criteria of CKD.

Secondly, previous genetic studies of CKD, both eGFRcr-driven and those based on several biochemical measures of CKD, have been hindered by a lack of human kidney tissue omics data encompassing the full regulatory cascade (from genome to proteome). Only with large scale epigenomics, transcriptomics and proteomics, in disease-relevant tissues, can we trace a causal path from genetic variant through epigenetic modifications, transcriptional regulation, protein production, to kidney dysfunction. Until very recently, proteomics – which provides critical information on the ultimate drivers of GWAS signals – has been the missing puzzle piece. Several studies integrating human genetics with plasma proteomics have proved a powerful strategy to dissect GWAS signals for different human disorders^13,15^. However, the use of disease-relevant tissue proteomics^16^ is much more limited, particularly in studies of cardiovascular and renal diseases – owing to the scarcity of human kidney datasets. Whilst two recent projects have leveraged kidney proteomics to identify protein drivers of CKD and blood pressure^10,17^, further integrative efforts are required to understand the cascades of molecular events, which are triggered by GWAS variants and culminate in CKD-predisposition.

Finally, whilst human genetics has long turned to monogenic disease genes for guidance in the search for common variant associations with disease^18–20^, the reverse analysis – in which symptoms and risks of rare monogenic disorders are suggested by GWAS of common variants – is uncommon. It is now becoming clear that many rare monogenic disease-causing variants are traceable in large genetic datasets^21,22^, and that common and rare variants in the same gene often exert similar phenotypic effects – albeit at different magnitudes of effect, and typically acting through different molecular mechanisms (*i.e.* common regulatory variants, rare coding variants)^17,23^. These developments facilitate a novel approach to the validation of GWAS drivers which, analogously to gene knock-out experiments, involves testing rare loss-of-function gene variants for phenotypic and disease associations in healthy populations.

Herein, leveraging data from up to 890,000 European-ancestry individuals from UK Biobank and CKDGen, we conducted GWAS meta-analyses of eGFRcr, eGFRcys, BUN and UACR, and identified 508 unique genetic locus regions associated with one or more CKD-defining traits. We integrated these with kidney tissue transcriptomics, proteomics and epigenomics data from up to 645 healthy kidneys curated from the Human Kidney Tissue Resource (HKTR), The Cancer Genome Atlas (TCGA) and Centre for Proteomic Tumour Analysis Consortium (CPTAC) to uncover 651 molecular partners, operating in kidney tissue, for 320 GWAS locus regions. Through the input from genome, kidney transcriptome, proteome and epigenome, we prioritized molecular targets for CKD, that act within the kidney, based on the body of existing evidence. Finally, we illustrate how a new high priority and clinically relevant kidney function gene - carbonic anhydrase 12 (*CA12*) - operates across the rare-common allele spectrum to determine susceptibility to renal and metabolic dysfunction.

## Results

### Genome-wide association study of four CKD-defining traits reveals overlapping genetic architecture

We performed autosomal GWAS of four CKD defining traits in UK biobank (UKBB) participants of European ancestry (eGFRcr, eGFRcys, BUN and UACR). To power our analyses, we meta-analysed the outputs from UKBB with the results of GWAS of synonymous traits conducted in participants of European ancestry from the CKDGen Consortium^3–5^, resulting in overall sample sizes of 888,929, 345,669, 553,039 and 438,658 respectively. We identified 68,690 (eGFRcr), 29,979 (eGFRcys), 19,092 (BUN) and 1,740 (UACR) genome-wide significant variants (*P* = 5×10^-8^, minor allele frequency – MAF > 0.5%). These corresponded to 421, 235, 198 and 51 unique loci, respectively – identified by a sliding window approach (**Fig. 1A-1B, Supplementary Tables 1-4, Methods**).

**Fig. 1:**
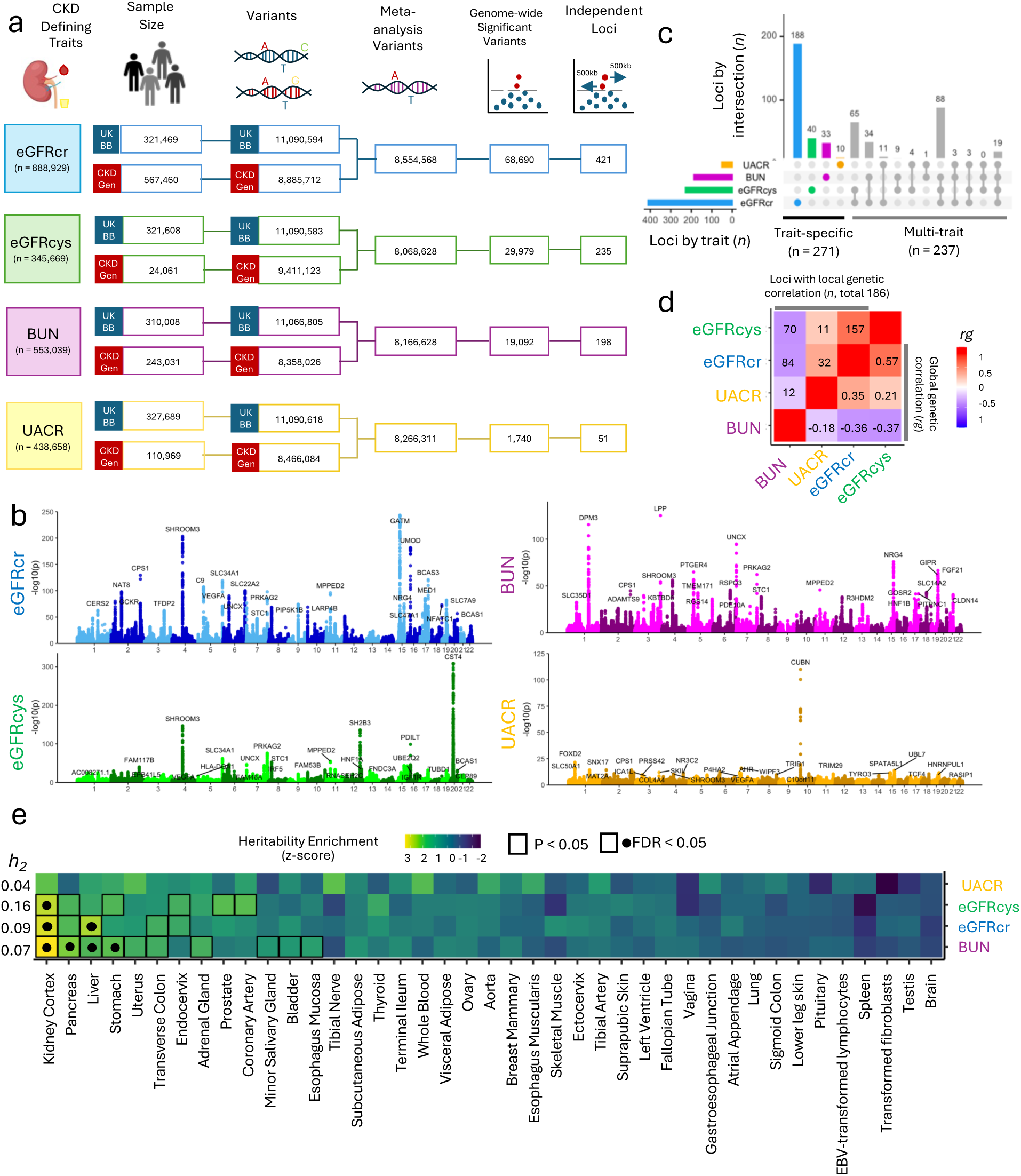
GWAS meta-analysis of 4 CKD-defining traits – unique and shared genetic signals. **A.** GWAS meta-analysis cohorts, sample sizes and numbers of tested variants, significant variants and independent locus regions. **B.** GWAS associations for each CKD-defining trait, *x* axis shows chromosomal position for 22 autosomes and *y* axis shows −log_10_P values for tested variants. For each GWAS variant, the 25 most significant loci are annotated by the gene nearest to the lead variant. **C.** Shared and trait-specific associations at 508 unique CKD locus regions. The left-hand bar-plot shows the number of locus regions (*x* axis) associated with each of the 4 CKD-defining traits (*y* axis), and the right-hand bar-plot shows the number of locus regions (*y* axis) associated with each trait combination (*x* axis), indicated by the bubble-plot below. **D.** Heatmap of global genetic correlation *rg* values from LDSC between trait pairs (below diagonal), and number of locus regions with significant local genetic correlations from LAVA (*FDR* < 0.05, above diagonal). Heatmap is coloured by *rg* values, from −1 (blue) to 1 (red). **E.** Heatmap of enrichment (normalized as a z-score) of tissue-specific genes in trait heritability (indicated by h_2_), calculated by stratified-LDSC using 53 GTEx tissues. Squares indicate nominal significance (P < 0.05), and points indicate *FDR* < 0.05 across all traits. Heatmap is coloured by heritability enrichment z-score per trait, ranging from −2 (blue) to 3 (yellow). Brain z-score represents the median z-score across 13 brain regions.

Loci were merged into 508 non-overlapping genomic regions (‘CKD locus regions, 237 of which encompassed more than one trait (‘multi-trait CKD locus regions’), and 271 were trait-specific (**Fig. 1C, Supplementary Table 5)**. A total of 124 multi-trait CKD locus regions were shared between two, 94 with three traits, and 19 between all four kidney traits. We identified 19 locus regions sharing association with all 4 CKD-defining traits. *SHROOM3*, the cystatin gene cluster, and *CUBN* are exemplar of multi- and trait-specific CKD locus regions respectively (**Extended Data Fig. 1**).

To further examine the shared genetic architecture of the 4 CKD-defining traits we estimated their global and local correlations using Linkage Disequilibrium Score Regression (LDSC) and Local Analysis of [co]Variant Association (LAVA)^24,25^. We observed significant global genetic correlations between all trait pairs (**Supplementary Table 6**). The strongest genetic correlation was observed between both eGFR measures (*rg* = 0.57, 157 CKD locus regions). BUN was inversely correlated with eGFRcr (*rg* = −0.36, 84 CKD locus regions) and eGFRcys (*rg* = −0.37, 70 CKD locus regions), while UACR showed a greater correlation with eGFRcr (*rg* = 0.35, 32 CKD locus regions) compared to eGFRcys (*rg* = 0.21, 4 CKD locus regions) or BUN (*rg* = −0.18, 12 regions) (**Fig. 1D).**

LDSC SNP-based heritability (h^2^) analysis estimated an h2 of 9%, 16%, 7%, and 4% for eGFRcr, eGFRcys, BUN, and UACR, respectively, consistent with prior estimates^5^. Stratified LDSC, partitioning heritability by gene expression annotations from 53 GTEx tissues, revealed marked significant heritability enrichment in kidney-specific genes for BUN, eGFRcr, and eGFRcys at a false-discovery rate (*FDR*) < 0.05 (**Fig. 1E, Supplementary Table 8**). CKD-defining traits were additionally enriched for genes expressed in tissues with prior evidence of involvement of their biomarker synthesis and metabolism (e.g. eGFRcr – liver; BUN – liver, stomach, pancreas).

Altogether, our multi-trait GWAS highlighted the shared genetic architecture of CKD – almost half of the uncovered GWAS regions are shared by two or more CKD-defining traits intersecting at genes expressed in the kidney. This illustrates a potential of the strategy bringing together the input from GWAS of several CKD-defining traits with kidney tissue omics, to identify and prioritise the genes, molecules and pathways underpinning genetic predisposition to CKD.

### CKD-defining trait heritability is mediated by kidney DNA methylation, gene expression, and protein abundance

To identify molecular mediators of genetic predisposition to CKD, we mapped molecular *cis*-quantitative trait loci (molQTL) using matched genetics and omics data from kidney samples in the Human Kidney Tissue Resource (HKTR)^17,26^, supplemented with normal adjacent control kidney tissue samples from The Cancer Genome Atlas (TCGA)^27^ and the Clinical Proteomic Tumor Analysis Consortium (CPTAC)^28^ (**Supplementary Table 9**). To match the GWAS study populations, we selected participants of European ancestry. We first examined a catalogue of genetic variants for association with DNA methylation *N* = 366, mRNA expression (*n* = 645), protein abundance (*n* = 83), and miRNA expression (*n* = 409). This identified DNA methylation QTLs (mQTLs) for 87,594 CpGs, mRNA expression QTLs (eQTLs) for 8,605 genes, protein abundance QTLs (pQTLs) for 243 proteins, and miRNA expression QTLs (mirQTLs) for 166 miRNAs (**Fig. 2A, Supplementary Tables 10-13**).

**Fig. 2.**
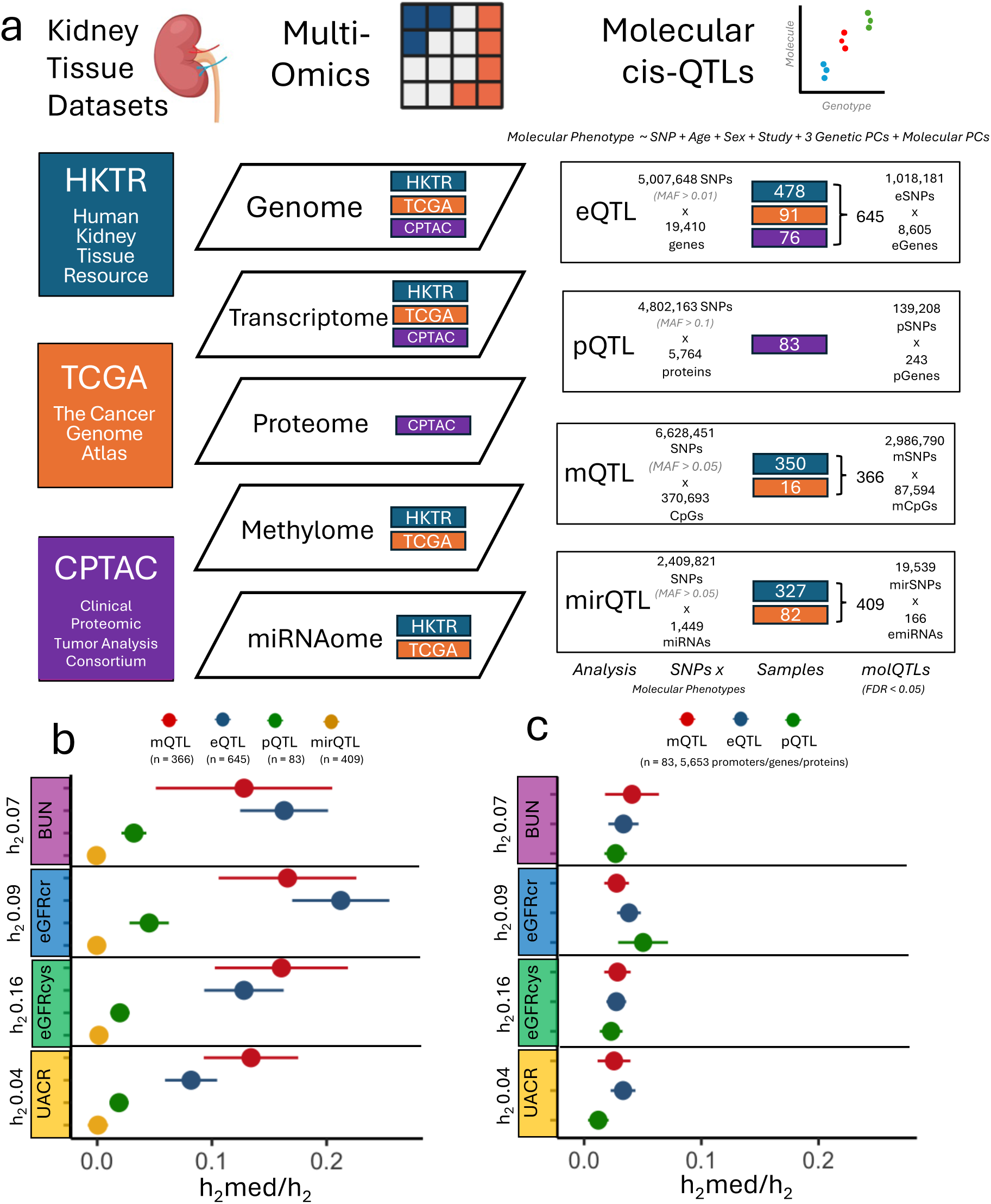
Integration analysis of CKD-defining traits GWAS with molQTL mapping in human kidney multi-omics. **A.** Sources of human kidney sample (left), omics data-layers (middle) and molQTL analyses (right). Coloured boxes indicate samples from each cohort (orange – HKTR, blue – TCGA, purple – CPTAC). **B.** Estimates of mediated heritability (h_2_med/h_2_) by molQTLs calculated by MESC. *X* axis shows h_2_med/h_2_ estimates and standard errors for each CKD-defining trait (*y* axis). Points are coloured by molQTL datatype (red – mQTL, green – eQTL, blue – pQTL, yellow – mirQTL), sample sizes are specified by *n*. **C.** Estimates of h_2_med/h_2_ by molQTL, downsampled to 83 random samples, for a common set of 5,653 genes, protein products and promoter CpGs. *X* axis shows h_2_med/h_2_ estimates and standard errors for each CKD-defining trait (*y* axis). Points are coloured by molQTL datatype (red – mQTL, green – eQTL, blue – pQTL, yellow – mirQTL).

We applied mediation expression score regression (MESC) to molQTL associations to estimate the proportion of trait heritability mediated by each omics layer (h2med/h2)^29^. Across the 4 CKD-defining traits, we found that mQTLs and eQTLs mediated similar proportions of heritability (mQTL: mean h2med/h_2_ =0.15, SE = 0.059; eQTL mean h_2_med/h_2_ =0.15, SE = 0.034), which was larger than that mediated by pQTLs (mean h^2^med/h^2^ =0.029, SE = 0.0011). mirQTLs mediated near zero heritability (mean h_2_med/h_2_ 0.00025, SE = 0.006) (**Fig. 2B).** We reasoned that pQTL-mediated heritability may be underestimated due to the small number of samples and proteins in the pQTL dataset. Therefore, we first down-sampled all molQTL datasets to 83 samples, and as expected, we found the heritability mediated by mQTLs and eQTLs reduced closer to the estimates for pQTLs – due to the relationship between h_2_med/h_2_ and sample size (**Extended Data Fig. 2A-B**). We then further equalized molQTLs to a uniform set of mRNAs, protein products, and promoter CpGs, and observed similar mediated heritability by mQTLs, eQTLs and pQTLs (**Fig. 2C**).

Compared to previous reports^11^ we observed similar mediation by eQTL and smaller mediation by mQTL (0.21 vs 0.46 mQTL h^2^med/h^2^). This was explained by the lower coverage of our analysis (this study: 370,693 CpGs, Liu et al.: 701,503 CpGs). Furtherer down-sampling by CpG number found that mQTL-mediated heritability increased with CpG coverage, suggesting that the heritability mediated by the full DNA methylome is underestimated (**Extended Data Fig. 2B**).

Collectively, our data demonstrate the importance of kidney DNA methylation, mRNA expression and proteins as mediators of CKD heritability.

### Colocalization between kidney eQTL and CKD-defining traits identifies novel and known disease-relevant gene

To investigate the transcriptional consequences of genetically determined kidney function, we leveraged our eQTL dataset generated with 645 kidney samples to examine the extent to which the expression of kidney genes colocalized with each CKD-defining trait. *Cis-eQTL* analysis identified renal genetic regulation for 8,605 of 19,410 tested genes (eGenes, 44%), by 1,018,181 variants (eSNPs, 20%), after hierarchical correction for multiple testing at an *FDR* of 0.05. To identify kidney eGenes which underpin genetic predisposition to CKD, we performed Bayesian colocalization analyses to identify eQTLs which share causal variants with CKD-defining traits. We identified 248 kidney eGenes which colocalized with one or more CKD-defining traits at a posterior probability (PP) > 0.8 **(Supplementary Table 14)**. These were termed ‘CKD-eGenes’. Of these, 189 genes colocalized with a single trait (trait-specific CKD-eGenes) and 59 were shared between multiple CKD-defining traits (multi-trait CKD-eGenes) (**Fig. 3A**).

**Fig. 3.**
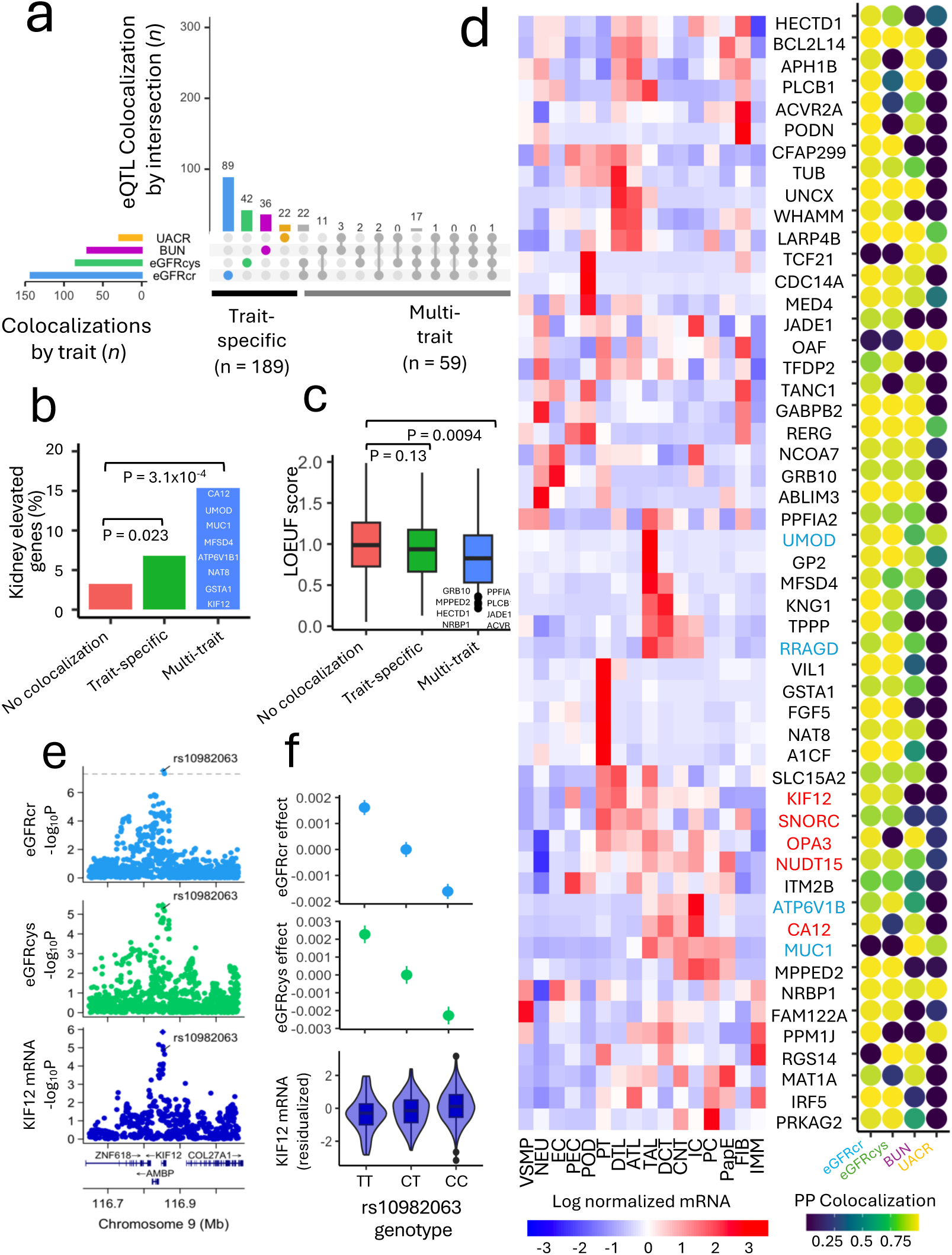
Colocalization between kidney eQTLs and multiple CKD-defining traits. **A.** Upset plot showing shared and trait-specific colocalizations for 248 eQTL signals which colocalized with one or more traits. The left-hand bar-plot shows the number of eQTLs (*x* axis) colocalizing with each of the 4 CKD-defining traits (*y* axis), and the right-hand bar-plot shows the number of eQTL (*y* axis) associated with each trait combination (*x* axis), indicated by the bubble-plot below. **B.** Proportion of kidney-enriched genes (*y* axis) in multi-trait eGenes, single-trait eGenes, and eGenes with no trait colocalizations (n = 5,733). *P* values from two-sided Fisher’s exact test. Boxplot showing distribution of LOEUF scores (*y* axis), where low scores indicate loss-of-function intolerance in multi-trait eGenes, single-trait eGenes and non-colocalizing eGenes. *P* values from two-sided Mann-Whitney U test. **D.** Left) Heatmap showing pseudobulk log normalized expression (z-score) of 46 protein-coding multi-trait eGenes for 16 cell-types. Heatmap is coloured by expression z-score, ranging from −3 (blue) to 3 (red). Gene names in red indicate ‘novel’ genes, which have not yet been suggested by CKD-GWAS gene prioritization studies. Gene names in blue indicate genes known to cause monogenic kidney disorders. Right) Bubbleplot showing GWAS-eQTL colocalizations for the 4 CKD-defining traits. Points are coloured by posterior probability of colocalization, ranging from 0 (blue) to 1 (yellow). **E.** LocusZoom plots for eGFRcr, eGFRcys and the *KIF12* eQTL, where the *x* axis shows chromosomal position, and the *y* axis shows −log_10_P values. Loci are annotated with the most likely causal variant **F.** Effect of rs10982063 genotype on eGFRcr, eGFRcys and renal *KIF12* expression. For eGFRcr and eGFRcys, the *y* axis gives the estimated effect of each additional allele on the residualized GWAS phenotype. Points show effect size estimates and lines show standard errors. For *KIF12* expression, the *y* axis shows the residualized rank-based inverse normal transformed and normalized mRNA abundance phenotype and the boxplots show the distribution of values for each genotype.

We hypothesized that genes colocalizing with multiple CKD-defining traits should show a stronger genetic relevance to kidney disease, than genes colocalizing with one trait alone^13^. To this end, we considered that ‘multi-trait CKD-eGenes’ would show both greater specificity to the kidney and greater constraint against damaging variation. Compared to non-colocalizing kidney eGenes (PP < 0.8 with all traits), multi-trait CKD-eGenes were significantly over-represented for kidney-enriched expression defined by Human Protein Atlas^30^ (odds ratio - OR = 5.4, *P* < 0.001, **Fig. 3B**), and were less tolerant of loss-of-function (LoF) variation, demonstrated by significantly lower average LOEUF (loss-of-function observed over expected upper fraction) scores^31^ (*P* = 0.0094, **Fig. 3C**). We observed numerically smaller over-representation of kidney-enriched expression (OR = 2.2, *P* = 0.03) and did not detect greater constraint (*P* = 0.31) for trait-specific CKD-eGenes when these were compared to non-colocalizing kidney eGenes (**Fig. 3C)**.

Given these findings, we focused further downstream annotations on multi-trait CKD-eGenes, as the strongest candidate drivers of genetic predisposition to CKD. We visualized 49 protein-coding multi-trait CKD-eGenes by their expression in 16 renal cell types, and annotated these by their colocalizing traits, involvement in monogenic kidney abnormalities, and novelty according to 16 prior studies^3–6,8–13,32–37^ (**Fig. 3D**, **Supplementary 15**). This showed that several of these genes have a strong prior relevance to monogenic kidney disorders (*UMOD*, *MUC1*, *ATP6V1B1*, *RRAGD*^38–40^) or were replicated by other CKD-GWAS (e.g. *MUC1*, *NAT8, IRF5*, *UNCX*^3,11,12^) while others were potentially new biologically plausible candidates. For example, kidney gene *KIF12*, encodes kinesin factor 12, a microtubule protein which is expressed in the developing kidney and adult kidney cell types^41^. *KIF12* is a modifier of polycystic kidney disease in mice^41,42^, and *KIF12* duplications in humans (likely increasing *KIF12* dosage) have been implicated in congenital abnormalities of the kidney and urinary tract in humans^43^. Through multi-trait colocalization^44^, we found that a shared causal variant was associated with higher *KIF12* mRNA, lower eGFRcr, and lower eGFRcys (PP = 0.96, **Fig. 3E-F**.)

Through these analyses, we found that the convergence of colocalization signals, between gene expression across several CKD-defining traits, reveals genes of greater biological relevance to CKD than the colocalization of single CKD-defining traits alone. This further underscores the importance of analysing multiple CKD-defining traits in the search for novel biologically relevant drivers of kidney function like *KIF12*.

### Integrative proteogenomic analysis of human kidney tissue reveals genetic mechanisms regulating the renal proteome

To explore the relationship between the genome, transcriptome and proteome in the kidney, we utilized the CPTAC proteomics dataset^28^, including 5,990 proteins quantified by tissue proteomics methodology (mass spectrometry) in 83 human kidney samples (**Extended Data Fig. 3A**).

First, we investigated the concordance of transcriptomic and proteomic measurements in the kidney for 5,973 proteins in 76 CPTAC samples of European ancestry with matching mRNA and protein measurements. By averaging across all samples, we found that median mRNA abundance was significantly correlated with median protein abundance (*r* = 0.47, *P* < 0.001, **Extended Data Fig. 3B**), and for each individual mRNA/protein pair we found that mRNA and protein measurements were weakly correlated on average (median *r* = 0.20, **Extended Data Fig. 3C**). This analysis also found a significant positive correlation (*FDR* < 0.05) for 2,407 (40%) kidney mRNA-protein pairs (**Supplementary Table 16**).

We next investigated the impact of common genetic variation on renal protein abundance by *cis*-pQTL analysis of 5,764 autosomally-encoded proteins using 83 available samples. We identified 14,634 genetic variants (pSNPs – 0.3%) regulating the abundance of 243 kidney proteins (pGenes, 4.2%) at *FDR* < 0.05. We compared the lead kidney pSNPs to lead kidney eSNPs (from our eQTL dataset) and found that lead kidney pSNPs were somewhat nearer to their respective TSS (median absolute distance 22kb *v* 27kb, inter-quartile range 7-63 *vs* 8-92kb, *P* = 0.065, **Extended Data Fig. 4A**). Furthermore, lead kidney pSNPs were partnered to significantly fewer pGenes than lead kidney eSNPs were to eGenes (range 1-2 *v* 1-14, *P* < 0.001, **Extended Data Fig. 4B**), however it is unclear whether this reflects greater specificity of protein regulation or is due to power.

We then examined the lead kidney pSNPs and pGenes through their molecular annotations. First, we annotated lead kidney pSNPs and non-pSNPs (variants with no association with any of the 243 pGenes) by functional scores, variant consequence, genomic context, and epigenomic modifications as well as chromatin states from ENCODE adult kidney tissue samples. Compared to non-pSNPs, lead kidney pSNPs were predicted to be more deleterious (median CADD Phred score 3.0 *vs* 2.4, *P* = 0.038) and more conserved across vertebrates (median phyloP 100-way score −0.064 *v* −0.27, *P* = 0.022, **Extended Data Fig. 4C**). Lead kidney pSNPs were also enriched for protein coding regions (missense, synonymous, exon, transcribed state), 5’ untranslated regions (UTRs), transcription factor binding sites (TFBSs), and gene promoters active in adult kidney tissue (H3K27Ac, H3K4Me3, CAGE (cap analysis of gene expression), chromHMM promoter state, **Extended Data Fig. 3D**).

We found that kidney pGenes had significantly greater median abundance (median log_2_ normalized abundance = 22.4 vs 21.0, *P* < 0.001), slightly greater median parent mRNA abundance (median log_2_ normalized abundance = 3.1 v 2.9, *P* = 0.083), and significantly greater mRNA-protein correlation than proteins without evidence of genetic regulation (median r = 0.43 v 0.19, *P* < 0.001, **Extended Data Fig. 4D**). We then performed gene-set over-representation analysis of biological processes (Gene Ontology) and found that metabolic and detoxification processes were over-represented in kidney pGenes (*FDR* < 0.01, **Extended Data Fig. 3E**, **Supplementary Table 17**). This replicates observations of kidney pGenes discovered using aptamer-based protein measurements^10^.

Finally, given that 84% of kidney pGenes (n = 203) were also kidney eGenes, we sought to quantify the extent of *cis*-eQTL support for *cis*-pQTL by: (i) eQTL-pQTL colocalization, and (ii) analysis of directionally consistent allelic effects (**Methods**). We uncovered that only 17% of kidney pQTLs colocalized with the eQTL signal for their parent gene (PP > 0.8, *N* = 42, **Extended Data Fig. 3F, Supplementary Table 18**), and that 62% of all lead pSNPs had directionally consistent eQTL effects (n = 150, **Extended Data Fig. 3G, Supplementary Table 19**). In contrast, <5% of kidney pQTL signals had directionally opposite eQTL effects (n = 9, **Extended Data Fig. 3G, Supplementary Table 19**), whilst the remaining 84 pQTL signals were not eQTLs. All colocalizing eQTL-pQTL pairs exhibited directionally consistent allelic effects apart from *DPP7*/DPP2, where the lead pQTL allele (rs11146000-T, predicted to disrupt a binding site for ETS-family TFs 247 bp upstream of the TSS) was associated with increased mRNA of the transcript but decreased abundance of the encoded protein in kidney tissue (eQTL effect = 0.40 and *P* = 7.1×10^-10^, pQTL effect = −1.0 and *P* = 3.4×10^-9^, PP = 0.94, **Extended Data Fig. 3G**).

To investigate the mechanisms of genetic regulation of kidney protein abundance, coupled to and uncoupled from the mRNAs, we compared functional annotations of lead kidney pSNPs from the pQTL analysis with (n = 150) and without eQTL support (n = 93 – includes both directionally opposite eQTL/pQTL and pQTL with no eQTL signal) to non-pSNPs. We found numerically greater enrichment of synonymous variants and regulatory annotations (TFBS, H3K27Ac, CAGE) in eQTL-supported kidney pSNPs, but greater enrichment of missense variants in kidney pSNPs without eQTL support (**Extended Data Fig. 4E**).

Collectively, our results point to the differences in genetic mechanisms of transcriptional and post-transcriptional regulation operating in the kidney. In line with other studies, of the kidney^10^ and other human tissues^45,46^, our data show that tissue protein levels only weakly correlate with their parent mRNA levels, and that more than a third of kidney pQTL are not supported by kidney eQTL signals. This further highlights the importance of pQTL analysis of disease-relevant tissues in the full functional appraisal of GWAS signals^10,17^.

### Integration of GWAS with kidney proteomics enhances gene prioritization for CKD-defining traits

To identify kidney protein drivers of CKD (hereafter CKD-pGenes), we integrated kidney proteomics data with our GWAS outputs. We nominated 50 CKD-pGenes based on: (i) Bayesian colocalization between kidney pQTLs and CKD-defining trait GWAS (PP H4 > 0.8, **Supplementary Table 14**), and/or (ii) directionally consistent allelic effects between kidney pQTLs and eQTLs (for CKD-eGenes), and/or (iii) a significant positive correlation between mRNA and protein abundance (for CKD-eGenes), (**Fig. 4A, Supplementary Table 20, Supplementary Note 1**).

**Fig. 4.**
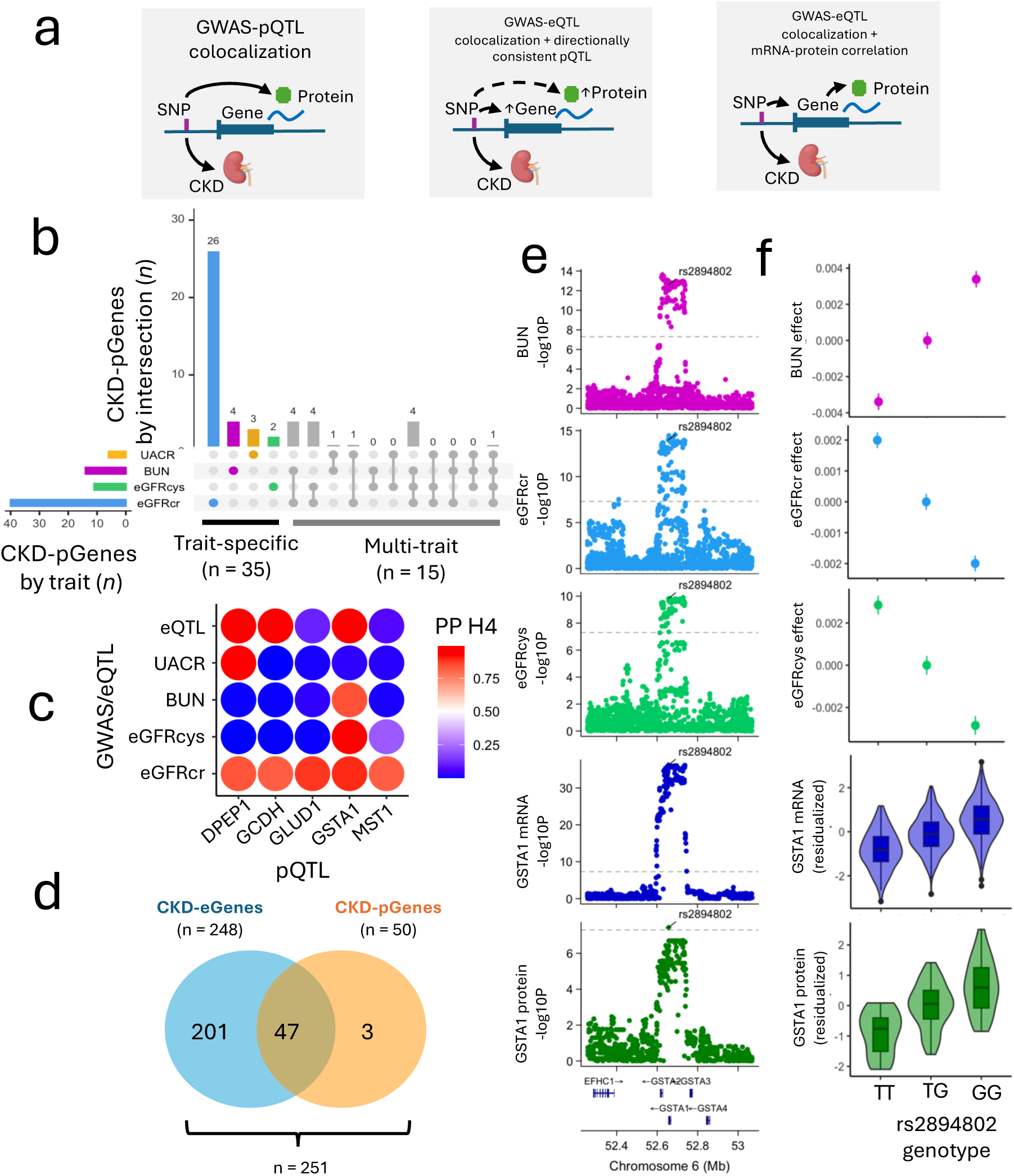
Integration of kidney tissue proteomics and GWAS to prioritize CKD-relevant proteins. **A.** Strategy to identify kidney proteins associated with CKD-defining traits (i.e. CKD-pGenes). CKD-pGenes were nominated by GWAS-pQTL colocalization, or GWAS-eQTL colocalization alongside directionally consistent pQTL, or significant kidney mRNA-protein correlation. **B.** Upset plot showing shared and trait-specific CKD-pGenes (n = 50). The left-hand bar-plot shows the number of pGenes (*x* axis) linked to each of the 4 CKD-defining traits (*y* axis), and the right-hand bar-plot shows the number of CKD-pGenes (*y* axis) associated with each trait combination (*x* axis), indicated by the bubble-plot below. **C.** Bubbleplot showing GWAS and eQTL colocalizations for the 5 CKD-pGenes nominated by kidney pQTL-GWAS colocalization. Points are coloured by posterior probability of colocalization (PP H4), ranging from 0 (blue) to 1 (red). **D.** Overlap between CKD-eGenes and CKD-pGenes. 248 CKD-eGenes were identified by colocalization between kidney eQTLs and CKD GWAS. 50 CKD-pGenes were identified by pQTL-GWAS colocalization, or directionally consistent eQTL-pQTL effects (for CKD-eGenes), or significant mRNA-protein correlation for (CKD-eGenes). The intersection of CKD-eGenes and CKD-pGenes is 47, and the union is 251. **E.** LocusZoom plots for BUN, eGFRcr, eGFRcys and *GSTA1* eQTL and pQTL signals, where the *x* axis shows chromosomal position, and the *y* axis shows −log_10_P values. Loci are annotated with the most likely causal variant **F.** Effect of rs2894802 genotype on BUN, eGFRcr, eGFRcys and *GSTA1* mRNA and protein expression. For BUN, eGFRcr and eGFRcys, the *y* axis gives the estimated effect of each additional allele on the residualized GWAS phenotype. Points show effect size estimates and lines show standard errors. For BUN mRNA and protein, the *y* axis shows the residualized mRNA/protein abundance phenotype and the boxplots show the distribution of values for each genotype.

Of these 50 CKD-pGenes, 35 showed pQTL or eQTL colocalizations with a single trait (trait-specific CKD-pGenes) while 15 pGenes demonstrated eQTL or pQTL colocalizations with multiple traits (multi-trait CKD-pGenes, **Fig. 4B, Supplementary**

**Table 20**). Whilst most CKD-pGenes were nominated by protein support from eQTL colocalization, five CKD-pGenes had evidence of pQTL colocalizations with CKD-defining traits - GCDH, GLUD1, and MST1 colocalized with eGFRcr only, whilst GSTA1 and DPEP1 exhibited multi-trait pQTL colocalizations (**Fig. 4C**).

We next examined the overlap between 50 CKD-pGenes and 248 CKD-eGenes identified earlier. This revealed 223 genes supported solely by mRNA colocalization, three supported exclusively by protein colocalization (*GCDH*, *GLUD1*, *MST1*), and 47 supported by both mRNA and protein (**Fig. 4D**). Among these, *GSTA1* emerged as a key example of a multi-trait CKD-pGene supported at both mRNA and protein levels. *GSTA1* encodes glutathione-S-transferase alpha 1, an enzyme involved in the detoxification of electrophilic compounds in liver and kidney proximal tubule. Notably, glutathione-S-transferase alpha enzymes are considered biomarkers of acute kidney injury (AKI), and *GSTA1* mRNA and protein are upregulated in mouse models of AKI ^47,48^. Using multi-trait colocalization, we identified a shared genetic regulation linking increased *GSTA1* mRNA and protein abundance with reduced kidney function, as measured by BUN, eGFRcr and eGFRcys (PP = 0.97, **Fig. 4E–F).** Importantly, no evidence of colocalization was observed for neighboring GST-alpha family members (*GSTA2*, *GSTA4,* **Supplementary Table 14**), suggesting that the association signal is specific to GSTA1. Collectively, these findings indicate that the genetic signal of association tracks through *GSTA1* mRNA and its protein product to culminate in changes to kidney function, irrespective of the biochemical test used to assess it.

In summary, our kidney proteogenomic analyses of CKD-defining traits identified disease-associated variants which operate through the renal expression of proteins. Importantly, by integrating proteomics data with the outputs from GWAS analyses, we identified and exemplified the full traction of CKD GWAS signals through several molecular layers of the kidney illustrating the benefit of integrative kidney multi-omics to post-GWAS investigations.

### Integration of GWAS, kidney DNA methylation, mRNA and protein expression identifies CKD-associated DNA methylation and kidney target genes

We next investigated genetically determined patterns of kidney DNA methylation associated with CKD defining traits, then characterized their downstream effects on kidney gene expression. To this end, we identified colocalizations between kidney *cis*-mQTLs and GWAS of CKD-defining traits and then linked colocalizing CpGs (CKD-mCpGs) to putative target genes (CKD-mGenes) by positional mapping and expression quantitative trait methylation (eQTM) analysis (**Fig. 5A**).

**Fig. 5.**
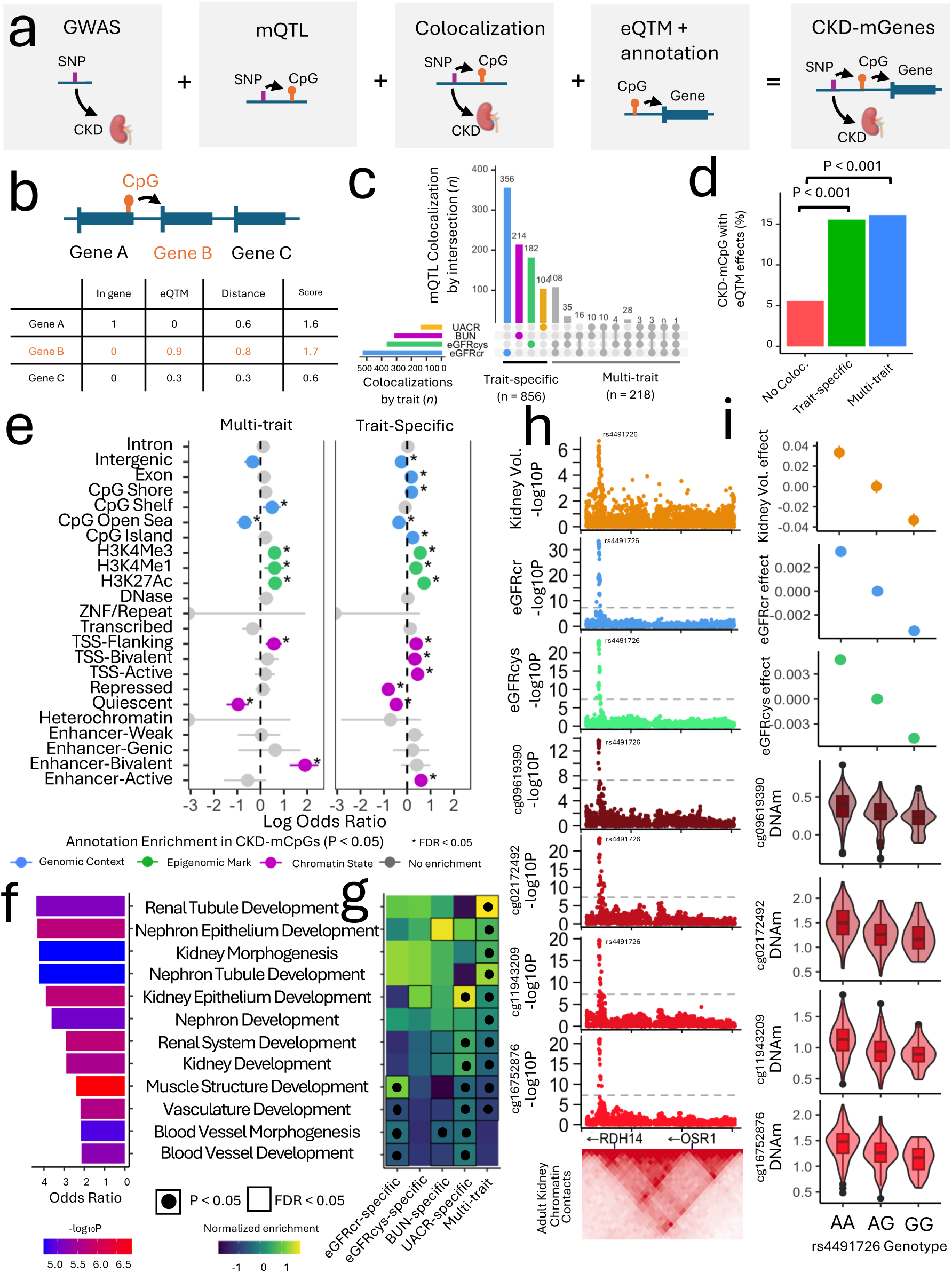
Colocalization between CKD-defining traits and kidney DNA-methylation. **A.** Strategy to identify genes targeted by DNA-methylation associated with CKD-defining traits. **B.** Examples of mCpG-mGene scoring. mCpGs are assigned to all TSSs < 1Mb, and scored by: (i) genic annotation, (ii) absolute beta coefficient of eQTM effect (*FDR* < 0.05) and (iii) TSS-CpG proximity. Each mCpG is assigned to the gene with the highest score (in this example gene B). **C.** Shared and trait-specific colocalizations (PP > 0.8) between 1,074 kidney mQTLs and one or more CKD traits. The left-hand bar-plot shows the number of mQTL (*x* axis) colocalizing with each of the 4 CKD-defining traits (*y* axis), and the right-hand bar-plot shows the number of mQTL (*y* axis) associated with each trait combination (*x* axis), indicated by the bubble-plot below. **D.** Enrichment of eQTM effects in trait-specific (n = 856) and multi-trait (n=218) CKD-mCpGs, compared to non-colocalizing mCpGs (n = 58,880). *X* axis shows proportion of CKD-mCpGs that overlap an emCpG. *P* values calculated by two-sided Fisher’s exact test. **E.** Enrichment of CpG annotations in trait-specific (n = 856) and multi-trait (n = 218) CKD-mCpGs, compared to non-colocalizing mCpGs (i.e. mCpGs with PP H4 < 0.8 for all traits, *N* = 58,880). *P* values calculated by a two-sided Fisher’s exact test. *X* axis shows log odds ratio (OR) for each annotation (*y* axis), where a log odds ratio > 1 indicates enrichment, and <1 indicates depletion. Points are coloured by annotation category, and lines show 95% confidence intervals. ‘*’ indicates significant enrichment correction for multiple testing (*FDR* < 0.05). **F.** Enrichment of gene-ontology biological processes in CKD-mGenes (n = 493), compared to non-CKD mGenes (i.e. non-overlapping genes assigned to mCpG with PP H4 > 0.8 for all traits, *N* = 10,192). *X* axis shows odds ratio for top enriched gene sets (*FDR* < 0.01), bars are coloured by −log_10_P value, calculated by one-sided Fisher’s exact test. **G.** Enrichment of the 12 significant gene sets in multi-trait CKD-mGenes and trait-specific CKD-mGenes, divided by trait (n: eGFRcr-specific − 157, eGFRcys-specific - 85, BUN-specific - 97, UACR-specific - 38, multi-trait - 116). Enrichment was calculated in comparison to non-CKD mGenes (n = 10,192). *P* values calculated by one-sided Fisher’s exact test. Colours of heatmap show odds ratio of enrichment from low (blue) to high (yellow). Outlined boxes indicate nominal (P < 0.05), and points indicate significance after correction for multiple testing (*FDR* < 0.05) **H.** LocusZoom plots for kidney volume, eGFRcr, eGFRcys, and kidney *cis*-mQTL for CKD-mCpGs (cg16752876, cg11943209, cg02172492 cg09619390) linked to OSR1. *X* axis shows chromosomal position, and *y* axis shows −log_10_P values. Loci are annotated with the most likely causal variant. Bottom shows heatmap of normalized chromatin contacts (low = white, high = red) between 50kb genomic bins from adult kidney Hi-C data. **I.** Effect of rs4491726 genotype on kidney volume, eGFRcr, eGFRcys and kidney DNA methylation of OSR1 CKD-mCpGs. For kidney volumes, eGFRcr and eGFRcys, the *y* axis gives the estimated effect of each additional effect allele on the residualized GWAS phenotype. Points show effect sizes and lines show standard errors. For the OSR1 CKD-mCpGs, the *y* axis shows the residualized DNA methylation phenotype, and the boxplots show the distribution of values for each genotype.

Using kidney mQTL data from 366 samples we detected *cis-*mQTLs and identified 87,594 CpGs (24% of 370,693) regulated by 2,986,790 genetic variants (45% of tested variants). We examined lead kidney mSNPs (n = 73,518) and found, on average, that each lead kidney mSNP regulated four mCpGs (range: 1–174). The median distance between lead mSNPs and their associated mCpGs was only 7,428 bp (IQR: 815 – 33,720bp). Compared to non-mSNPs, lead kidney mSNPs were significantly enriched for coding and regulatory annotations (**Extended Data Fig. 5A),** and preferentially overlapped kidney eSNPs (P < 0.001; **Extended Data Fig. 5B**) and kidney pSNPs (P < 0.001; **Extended Data Fig. 5C**).

To evaluate how kidney CpG methylation associates with gene expression, we performed a *cis*-eQTM analysis of the same 366 kidney samples. We identified 6,040 significant associations between 5,057 CpGs (emCpGs; 1.4% of 369,175 tested) and 1,761 kidney genes (emGenes; 9.0% of 19,427 tested, **Supplementary Table 21, Supplementary Note 1**). Kidney emCpGs were located a median of 17,922 bp from their target TSS (IQR: 2,009–70,782 bp) and typically regulated a single gene (range: 1–7). Most eQTMs exhibited inverse associations between DNA methylation and gene expression (negative eQTMs, *N* = 3,720); fewer had a directionally consistent relationship (positive eQTMs, *N* = 2,320), exceeding random expectation (*P* < 0.001, binomial test) (**Extended Data Fig. 5D**).

We observed that 1,792 emCpGs exhibited uniquely activating effects and that 3,048 exhibited uniquely repressive effects. 217 CpGs had both positive and negative associations with different emGenes. Notably, negative emCpGs were located closer to their target TSSs than positive emCpGs (median distance: 10,660 vs. 35,992 bp, IQR: 863–49,592 vs. 7,172–102,106 bp, respectively, **Extended Data Fig. 5D**), and were enriched for indicators of enhancer, and promoter-flanking chromatin (**Extended Data Fig. 5E**). In contrast, positive emCpGs, which tended to be more distal, were enriched for polycomb-repressed chromatin (**Extended Data Fig. 5D-E**). These observations are consistent with previous eQTM studies of the kidney and other tissues^11,49^.

We leveraged our kidney eQTM data to prioritize target genes for kidney mCpGs (i.e. kidney mGenes). Here we computed a prioritization score for candidate mCpG-gene pairs (CpG <1 Mb from TSS) incorporating: (i) mCpG position within a gene body, (ii) mCpG-TSS distance, and (iii) magnitude of kidney eQTM effect. Each kidney mCpG was then assigned to the gene with the greatest score (**Fig. 5B, Supplementary Table 22**).

By integrating data from GWAS of CKD-defining traits, kidney mQTL, and kidney eQTM we identified 1,074 CKD-mCpGs linked to 493 kidney mGenes. 1,074 CKD - mCpGs were nominated by kidney mQTL colocalizations with CKD-defining traits (PP > 0.8, **Supplementary Table 14, Supplementary Note 1**) and included 856 trait-specific CKD-mCpGs (which colocalized with a single trait) and 218 multi-trait CKD-mCpGs (which colocalized with multiple traits, **Fig. 5C**). Correspondingly, we defined 377 trait-specific CKD-mGenes (partnered with trait-specific CKD-mCpGs for the same trait) and 116 multi-trait CKD-mGenes (partnered with multi-trait CKD-mCpGs, or trait-specific CKD-mCpGs for different traits, **Supplementary Fig. 1, Supplementary Table 23**).

We next investigated how CKD-associated methylation patterns influence gene expression in the adult kidney. 168 CKD-mCpGs (16%) showed kidney eQTM effects on 81 genes (CKD-emCpGs and CKD-emGenes, **Supplementary Note 1**). Both multi-trait and trait-specific CKD-mCpGs were significantly enriched for eQTM effects compared to 58,880 CKD-non-colocalizing mCpGs (*P* < 0.001, **Fig. 5D**). We observed both negative and positive associations. For example, hypermethylation of the CKD-emCpG site cg06630958 in the *IRF5* promoter was linked to lower *IRF5* expression (*P* = 6.6×10^-8^, effect = −0.28, **Extended Data Fig. 6A**). In contrast, hypermethylation of another CKD-emCpG – cg06766541, located ∼21 kb upstream in a polycomb-repressed region – was associated with higher *IRF5* expression (*P* = 8.5×10^-9^, effect = 0.30, **Extended Data Fig. 6A**). We then overlapped CKD-mGenes (n = 493) with the union of CKD-eGenes and CKD-pGenes (*n* = 251). We found that CKD-emGenes (*n* = 51), compared to CKD-mGenes without eQTMs (*n* = 442), were enriched for genes nominated by kidney eQTL and pQTL colocalization (*n* = 442, OR = 14, *P* < 0.001, **Extended Data Fig. 6B**).

### Kidney DNA methylation at renal development genes colocalizes with multiple CKD GWAS traits

We further characterized 1,074 CKD-mCpGs by genomic context, alongside epigenomic modifications and chromatin states in adult kidney tissue. Compared to CKD non-colocalizing mCpGs (*n* = 58,880), both multi-trait and trait-specific CKD-mCpGs were depleted from intergenic and ‘open-sea’ CpG regions and enriched for epigenomic indicators of promoter and enhancer activity, alongside TSS-flanking chromatin (**Fig. 5E, Supplementary Table 22**). Interestingly, bivalent enhancers – defined by cooccurring activating and repressive histone marks and typically involved in developmental gene regulation^50^ – were only significantly enriched in multi-trait CKD-mCpGs (OR = 6.7, *FDR* < 0.001), but not in trait-specific CKD-mCpGs (*FDR* = 1, **Fig. 5E**).

To investigate the biological function of the 493 CKD-mGenes, we performed gene set over-representation analysis of GO biological pathways, using 10,192 mGenes partnered with non-colocalizing mCpGs as a reference. We found significant enrichment for biological processes related to kidney, muscle and vascular development (*FDR* < 0.05, **Fig. 5F, Supplementary Table 24**).

To investigate whether enrichment of these processes was driven by multi-trait or trait-specific CKD-mGenes, we identified the top 12 enriched gene sets (*FDR* < 0.01) and analyzed enrichment separately (and divided trait-specific CKD-mGenes by the colocalizing trait). This revealed distinct patterns: the muscle development gene set was preferentially enriched among eGFRcr-specific kidney mGenes, whilst kidney developmental gene sets were preferentially enriched among multi-trait CKD- mGenes (**Fig. 5G, Supplementary Table 25**). These included established embryonic and fetal nephrogenesis genes such as *BMP4*^51,52^*, OSR1*^53–56^*, OVOL1*^57^*, PAX8*^58,59^*, PRICKLE1*^60,61^*, TCF21*^62,63^ *and TFAP2A*^64,65^. We also observed enrichment of genes involved in kidney epithelium development in UACR-specific genes (**Fig. 5G, Supplementary Table 25**).

Given that CKD-mGenes were enriched for genes which primarily operate during organ development, we investigated whether variants influencing developmental gene methylation, also influenced gene expression in adult kidney. To answer this question, we focused on a set of 26 CKD-mGenes with established roles in renal system development (**Methods**) and investigated their partnered CKD-mSNPs and CKD-mCpGs (hereafter referred to as ‘developmental CKD-mGenes’, ‘developmental CKD-mCpGs’ and ‘developmental CKD-mSNPs’ respectively, **Supplementary Table 26, Supplementary Note 1**). We next tested the proportion of developmental CKD-mSNPs that were also eSNPs in adult kidney tissue. Developmental CKD-mSNPs (*n* = 42) resulted being significantly depleted of kidney eSNPs, compared to the remaining 552 CKD-mSNPs not linked to renal development genes (OR = 3.9, *P* < 0.001, **Extended Data Fig. 7A**), suggesting that developmental CKD-mQTL do not affect CKD-defining traits through modulating transcriptional levels in the adult kidney.

To further characterize molecular mechanisms of developmental CKD-mQTL we then explored functional annotations of developmental CKD-mCpGs. When compared to other ‘non-developmental’ CKD-mCpGs (*n* = 1,006), developmental CKD-mCpGs (*n* = 68) were uniquely enriched for bivalent promoters (*FDR* < 0.05) – which poise genes for activation at specific developmental timepoints^50^ **(Extended Data Fig. 7B)**.

Finally, given that developmental gene function was enriched among multi-trait CKD-mCpG, we wished to examine whether the developmental CKD-mCpG colocalized with multiple or single CKD-defining traits. Supporting our previous results, we found a significantly greater proportion of developmental CKD-mCpGs in multi-trait CKD-mCpGs (14%) than in trait-specific CKD-mCpGs (5%, *P* < 0.001, OR = 3.2, **Extended Data Fig. 7C)**.

To exemplify our findings, we investigated the developmental CKD-mQTL for *OSR1* – a novel candidate driver of CKD-defining trait GWAS. *OSR1* encodes the odd skipped-related 1 transcription factor, which is active in kidney organogenesis and associated with kidney developmental defects in mice^53,55^. The variant rs4491726-G allele was associated with lower eGFRcr, lower eGFRcys, and lower kidney DNA methylation at 4 CpG sites located in a polycomb-repressed region in *OSR1* exon 2 (**Fig. 5H-I, Supplementary Table 22**). Intriguingly, rs4491726 was located 863kb from *OSR1*, but connected by a long-range chromatin loop (**Fig. 5H-I**). Neither rs4491726, nor the *OSR1* CKD-mCpGs, were associated with adult kidney expression of *OSR1* (eQTL *P* = 0.19, eQTM *P* range = 0.11-0.46) or any other gene. Since *OSR1* variants have been suggested to cause kidney hypoplasia in humans^54,56^, we investigated GWAS of kidney volume in European UK Biobank participants^66^, and found that rs4491726-G was associated with reduced kidney volume (*P* = 7.7×10^-7^, **Fig. 5H-I**). Multi-trait colocalization confirmed a shared causal variant for reduced methylation of the polycomb-repressed *OSR1* element, smaller kidney volume and lower eGFR (PP = 0.99, **Fig. 5H-I)**.

In summary, our integrative analysis of CKD-defining trait GWAS and kidney DNA methylation, we identified two distinct mechanisms of CKD-associated DNA methylation: CKD-eQTMs and developmental CKD-mQTLs. The CKD-eQTMs were consistent with the canonical regulatory cascade in adult kidney tissue, where genetically determined increases in DNA methylation at promoters and enhancers were associated with transcriptional repression of CKD-mGenes (e.g. cg06630958 and *IRF5*). We further observed the inverse effect, with genetically increased DNA methylation at polycomb-repressed elements (i.e., *cis*-regulatory silencers^67^) associated with upregulated expression of the CKD-mGene, exemplified by cg06766541 and *IRF5*. CKD-mGenes nominated by kidney eQTMs strongly overlapped with CKD-eGenes and CKD-pGenes.

The second mechanism of CKD-associated DNA methylation records the regulation of kidney development genes (developmental CKD-mQTLs) and is exemplified by *OSR1*. We found that the constituent developmental CKD-mSNPs typically had no effect on adult kidney transcription, and their partnered developmental CKD-mCpGs were preferentially located in bivalent chromatin structures at developmental gene promoters. These properties of developmental CKD-mQTL suggest that these genetic variants, which influence CKD-defining traits, may act by modulating embryonic or fetal gene expression within the developing kidney. Our findings are consistent with evidence that cells retain an ‘epigenetic memory’ of developmental gene regulation – in which the methylation marks deposited during embryonic/fetal gene regulation are retained across cell divisions^68,69^. Importantly, we found that developmental CKD-mQTLs were enriched for colocalizations with multiple CKD-defining traits. Altogether these findings suggest that genetic regulation of renal development is: (i) recorded in the methylomes of adult kidney cells, and (ii) influences multiple measures of kidney health in later life.

### Multi-omic, multi-trait gene prioritization illuminates targetable genes, molecules, and pathways underlying genetic predisposition to CKD

We collated 248 CKD-eGenes, 50 CKD-pGenes, and 493 CKD-mGenes from our earlier integrative analyses to nominate 651 unique kidney genes as candidate drivers of CKD-defining trait GWAS. We termed these ‘CKD kidney genes’, **Supplementary Note 1**. To further prioritize these targets, we devised an intuitive gene prioritization strategy where CKD kidney genes were scored based on the cumulative support across four CKD-defining traits and three kidney omics layers. Here, each CKD kidney gene was scored from 0-12 by the number of trait associations on the eGene (0-4), pGene (0-4), and mGene (0-4) level, following the definitions outlined previously (**Fig. 6A-B, Supplementary Table 27**). CKD kidney genes covered 320 (62%) of the 508 unique GWAS locus regions, with a median of 1 gene nominated per region (range 0-19). CKD kidney genes covered significantly more multi-trait locus regions (75% of 237) than trait-specific regions (53% of 271, *P* < 0.001, **Fig. 6C, Supplementary Table 28**).

**Fig. 6.**
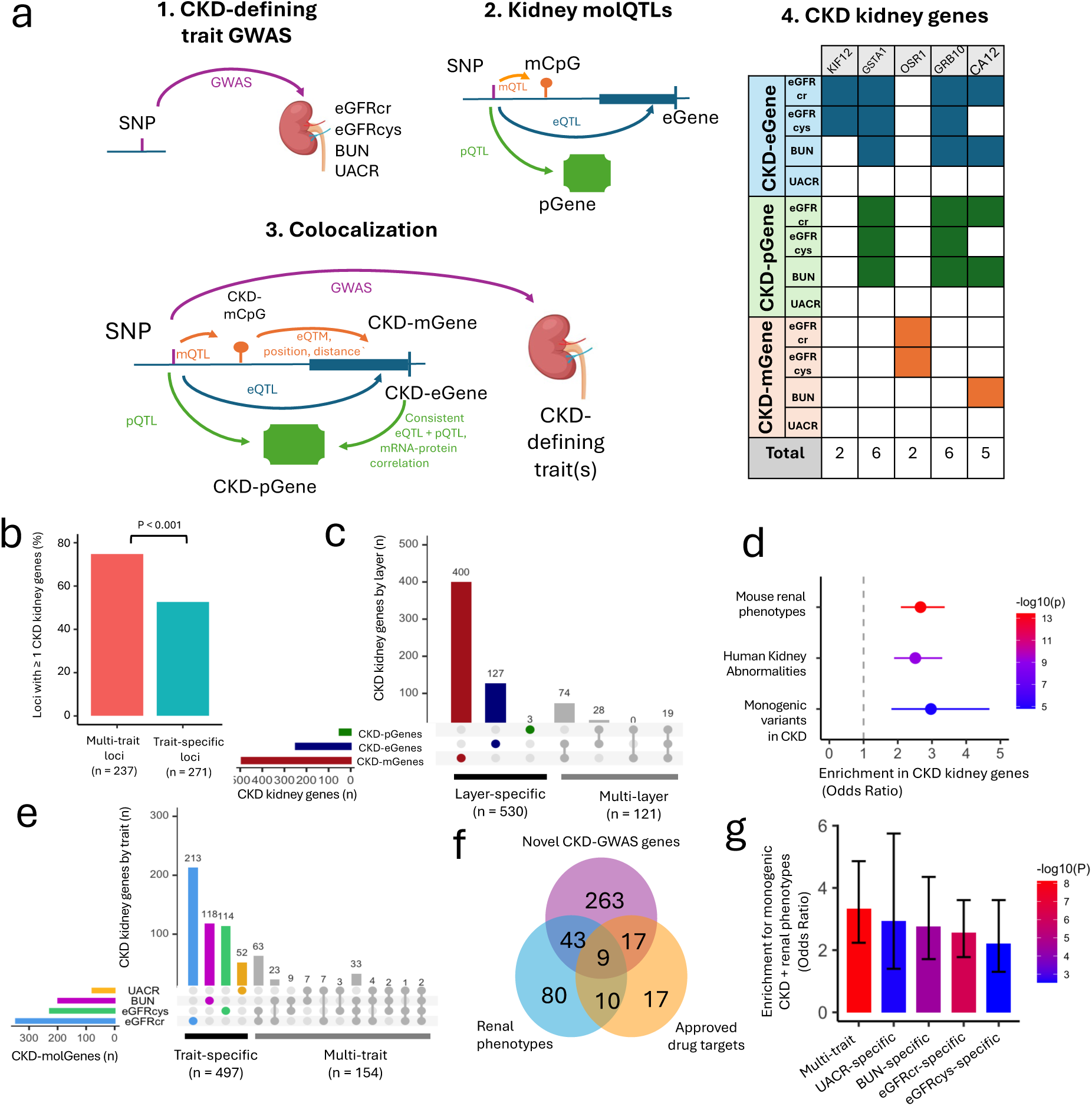
Nomination and prioritization of CKD kidney genes. **A.** Schematic showing the nomination and prioritization of CKD-kidney genes at CKD-defining trait locus regions. Genetic locus regions associated with CKD-defining traits are first identified (1), followed by discovery of molQTLs in human kidney tissue (2). Kidney genes are then linked to CKD-defining traits through the colocalization of CpGs, genes and proteins (3). Finally, 651 nominated genes are scored by the number of association on each molecular level, out of a maximum score of 12 (representing 4 trait colocalizations with 3 molecular layers) **B.** Percentage of locus regions where at least one CKD kidney gene was nominated as a causal GWAS gene (*y* axis) for multi-trait and trait-specific locus regions (*x* axis). **C.** Upset plot showing 651 CKD kidney genes prioritized across each molecular layer. The left-hand bar-plot shows the number of CKD kidney genes (*x* axis) supported by each layer (*y* axis), and the right-hand bar-plot shows the number of CKD kidney genes (*y* axis) with support from each combination of layers (*x* axis), indicated by the bubble-plot below. **D.** Enrichment of single gene causes of CKD and renal abnormalities (in mice and humans) within protein-coding CKD kidney genes. *Y* axis shows three renal phenotype gene sets tested for enrichment in 651 CKD kidney genes, and X axis shows the odds ratio (points) and 95% CIs (lines) of enrichment in comparison to 9,305 neighbouring genes within matched GWAS locus regions. Points are scaled by the number of CKD kidney genes within the gene set and coloured by −log10Pvalues, calculated by two-sided Fisher’s exact tests. N-values indicate the number of CKD kidney genes within each gene set. **E.** Upset plot showing shared and trait-specific CKD kidney genes. The left-hand bar-plot shows the number of CKD kidney genes (*x* axis) prioritized for each of the 4 CKD-defining traits (*y* axis), and the right-hand bar-plot shows the number of CKD kidney genes (*y* axis) associated with each trait combination (*x* axis), indicated by the bubble-plot below. **F.** Overlap of CKD kidney genes annotated by novelty (n = 330), renal abnormalities/diseases (n = 142), and FDA-approved drug targets (n = 53). **G.** Enrichment of renal phenotype genes in CKD kidney genes divided into multi-trait (n = 154), and trait-specific genes (n: eGFRcr – 213, eGFRcys – 113, BUN – 118, UACR - 52). *Y* axis shows the odds ratio for enrichment of renal phenotypes within each gene set (*x* axis). Bars are coloured by −log_10_ *P* value calculated by two-sided Fisher’s exact tests against the set of 9,305 neighbouring genes that were not prioritized. Error bars indicate 95% confidence intervals.

To investigate the relevance of our prioritized GWAS targets to kidney disease, we compared the 651 CKD kidney genes to a set of neighbouring protein-coding and lncRNA genes located within the same set of 320 GWAS locus regions (*n* = 9,035). We found that CKD kidney genes were enriched for renal abnormalities in mouse knock-out models (Mouse Genome Informatics^70^), genetic causes of human renal abnormalities (Human Phenotype Ontology^71^), and genetic causes of human CKD (Groopman et al. 2019^72^, *P* < 0.001, **Fig. 6D**). We then pooled these annotations to generate a set of single gene causes of kidney abnormalities/diseases from the listed sources (**Supplementary Table 29**), and tested their enrichment in our CKD kidney genes, divided by multi-trait and trait-specific genes (**Fig. 6E**). We found that whilst all groups were enriched, compared to the 9,035 background genes, multi-trait CKD kidney genes exhibited the greatest level of enrichment for renal abnormalities/diseases (**Fig. 6G**). This finding further demonstrates that kidney genes prioritized by multiple CKD-defining traits best capture the underlying genetic susceptibility to CKD.

We then annotated our CKD kidney genes by novelty, prior relevance to human disease and drugs and found: 142 genes associated with CKD or kidney phenotypes in humans or mice^70–72^; 330 novel drivers of CKD-defining trait GWAS (not prioritized by previous studies^3–6,8–13,32–37^); and 53 targets of approved or investigational therapies from Open Targets^73^ – which may present opportunities for drug repurposing (**Fig. 6F, Supplementary Table 30**). A scored table of CKD kidney genes, alongside their novelty, associated renal phenotypes and current therapeutic status is provided in **Supplementary Table 27** as a resource to inform follow-up studies. We further performed in depth functional annotations of the top scoring 40 genes (score > 4) against the current literature, showing their known molecular, cellular and biological functions alongside any known relevance to kidney function or disease (**Supplementary Table 31**). These annotations revealed additional CKD kidney genes with potential therapeutic relevance, including *GRB10*.

*GRB10* encodes growth factor binding protein 10, an adapter protein which modulates insulin and insulin-like growth factor signalling^74,75^. GRB10 has been implicated in the amelioration of diabetic nephropathy by catalpol^76^– an active compound in the Chinese traditional herbal medicine *Rehmannia glutinosa* **(Extended Data Fig. 8A).** In a multi-centre randomized control trial *Rehmannia*, improved kidney function in type 2 diabetics with CKD^77^, and evidence from animal studies suggests that catalpol itself is protective against kidney damage induced by high-glucose levels^78^. We found that the genetically determined decrease in kidney expression of *GRB10* gene correlates with its protein abundance in the kidney and colocalized with higher eGFRcr, higher eGFRcys and lower BUN. The direction of these associations is consistent with a previously demonstrated downregulation of GRB10 promoting the improvement in kidney function upon catalpol treatment of mice (**Extended Data Fig. 8A**)^76^. HyPrColoc confirmed a shared causal variant between decreased renal *GRB10* and increased kidney function measured by three biochemical indices (PP = 0.99, **Extended Data Fig. 8B-C**).

Given the known roles of GRB10 in insulin signalling, growth, and other health-relevant processes, we then investigated phenome-wide associations with kidney *GRB10* mRNA. Using our previous strategy^17^, we tested genetically-predicted kidney *GRB10* expression for association with 193 quantitative traits in UK Biobank (*n* = 337,350, European ancestry), revealing associations with 18 phenotypes (*FDR* < 0.05). Amongst these were kidney function, serum urate, and blood glucose levels (**Supplementary Table 32**). These results show that the genetically determined reduction of kidney *GRB10* has pleiotropic effects on human health including increased kidney function, decreased blood glucose and lower levels of urate.

Altogether integration of input from three layers of kidney omics when combined with the data from GWAS of multiple CKD-defining traits can help prioritising hundreds of potential contributors to CKD. We also exemplify how this information can shed further light on the biological underpinnings of the uncovered associations and/or selection of potentially actionable targets for future studies.

### Genetically-determined reduction of renal carbonic anhydrase 12 is associated with decreased kidney function and increased serum urate

Our analyses prioritised *CA12* as a novel CKD kidney gene targeted by existing therapeutics. *CA12* encodes a carbonic anhydrase (CA) enzyme, which catalyzes the reversible hydration of carbon dioxide to bicarbonate and protons. CA12 is expressed on the basolateral membrane of kidney tubular cells, where it functions to maintain pH and electrolyte balance (**Fig. 7A**). CA12 (alongside CA2 and CA4) is inhibited by acetazolamide^79^ – a weak diuretic promoting the excretion of electrolytes in alkalinized urine (**Fig. 7B**).

**Fig. 7.**
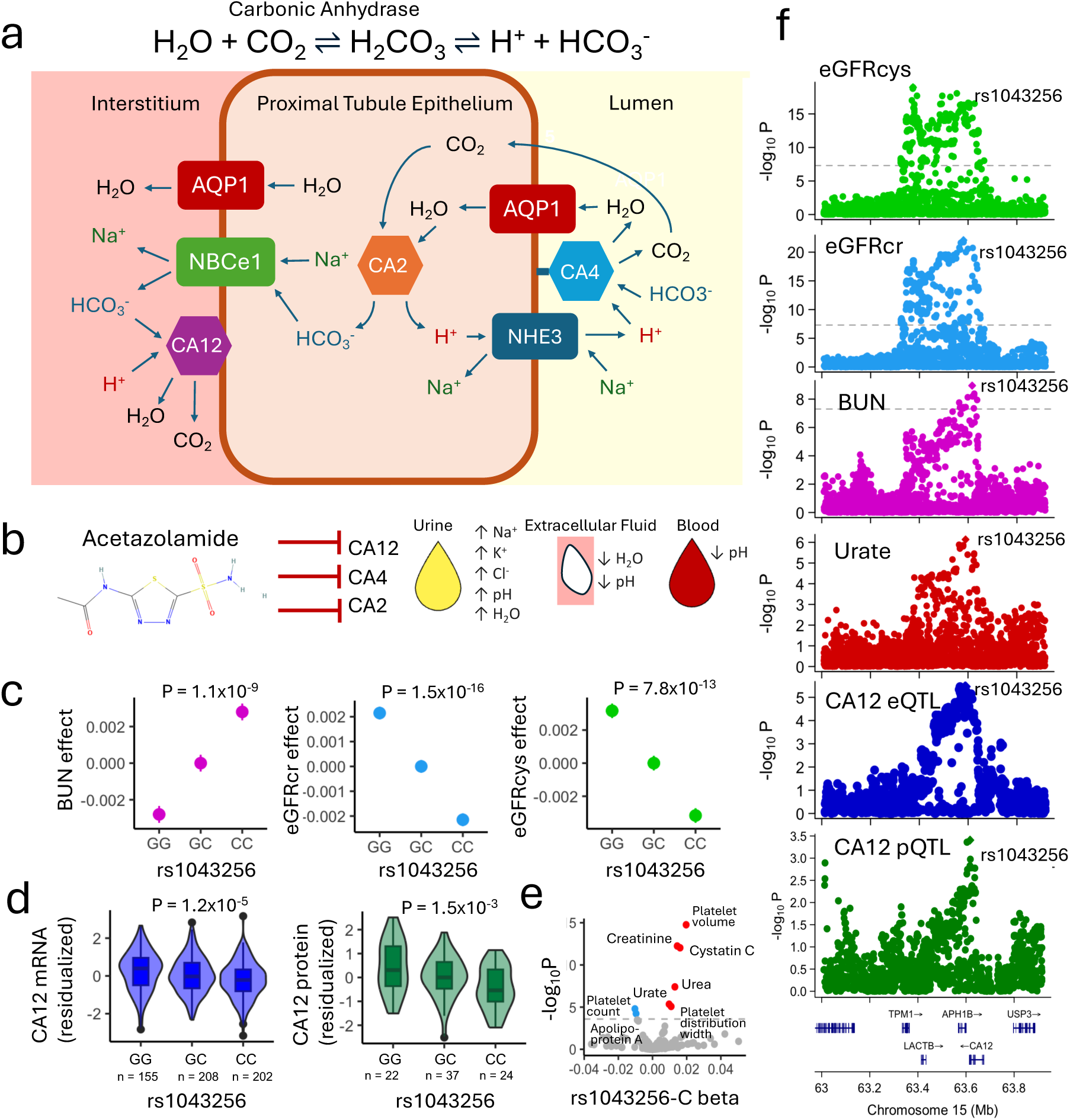
Shared causal genetic associations between reduced kidney CA12 levels, decreased kidney function, and increased serum urate. **A.** Enzymatic activity of CA12 in the kidney proximal tubule. CA12 activity is shown in context with other renal enzymes and transporters to demonstrate how the push-and-pull of bicarbonate and protons maintains pH and sodium balance. **B.** Inhibition of CA12, alongside CA2 and CA4 by acetazolamide treatment, and its effects on pH, electrolyte balance, and fluid volume in urine, blood and extracellular fluid. **C.** Effects of rs1043256 on eGFRcr (N = 888,929), eGFRcys (N = 345,669), and BUN (N= 555,039) in the UKBB-CKDGen GWAS meta-analysis. Effect size and standard errors are shown on the *y* axis for each copy of the C allele on the *x* axis. **D.** Effects of rs1043256 on *CA12* mRNA and protein identified by eQTL (N = 645) and pQTL (N = 83) analysis in human kidney. Distributions of *CA12* mRNA (residualized RBINT normalized log_2_ TPM) and protein (Residualized log_2_ normalized protein abundance) are shown on the *y* axis for each rs1043256 genotype on the *x* axis. **E.** Effects of rs1043256 on 193 quantitative phenotypes in European UKBB participants (N ≤ 336,950). Volcano plot shows −log10P values on y axis, and effect size of rs1043256 C allele on *x* axis. Significant positive associations (below the Bonferroni-adjusted threshold *P* < 2.6×10^-4^) are shown in red, and significant negative associations shown in blue. **F.** GWAS and QTL associations for variants at CA12 locus. LocusZoom plots show −log10P values on x axis and genomic position on *y* axis for all tested variant associations within the CA12 locus.

Our results identified that common genetic variants (including rs1043256-C in the *CA12* 5’UTR) associated with reduced kidney *CA12* mRNA and protein, were also associated with reduced kidney function measured by eGFRcr, eGFRcys and BUN (**Fig. 7C-8D)**. Further phenome-wide association study (PheWAS) of rs1043256 in up to 336,950 UKBB participants of European ancestry identified that its C allele also increased serum urate levels (**Fig. 7E**). Our multi-trait colocalization using HyPrColoc revealed that all these associations were driven by the same causal variant (**Fig. 7F**). This analysis examined *CA12* kidney *cis*-eQTL and *cis*-pQTL associations, alongside summary statistics for eGFRcr, eGFRcys, BUN and serum urate^80^. We first identified multi-trait colocalizations between eGFRcr, BUN, urate and *CA12* mRNA (PP = 0.98, **Fig. 7F**). We found no colocalization between *CA12* expression and eGFRcys but noticed the presence of an additional signal near known kidney function gene - *LACTB*^81^. To separate out these two signals, we conditioned eGFRcys associations on the lead variant and performed multi-trait colocalization using the conditioned eGFRcys associations. We then found evidence that *CA12* expression, eGFRcr, eGFRcys, BUN and urate shared a causal variant (PP = 0.93, **Fig. 7F.)**. Sensitivity analyses showed that colocalization of the weaker CA12 pQTL signal was sensitive to HyPrColoc parameter choice (**Supplementary Fig. 2**). Collectively, these data indicate that rs1043256 (or a co-inherited variant) modulates both renal function and urate levels by altering renal expression of *CA12*.

**Fig. 8.**
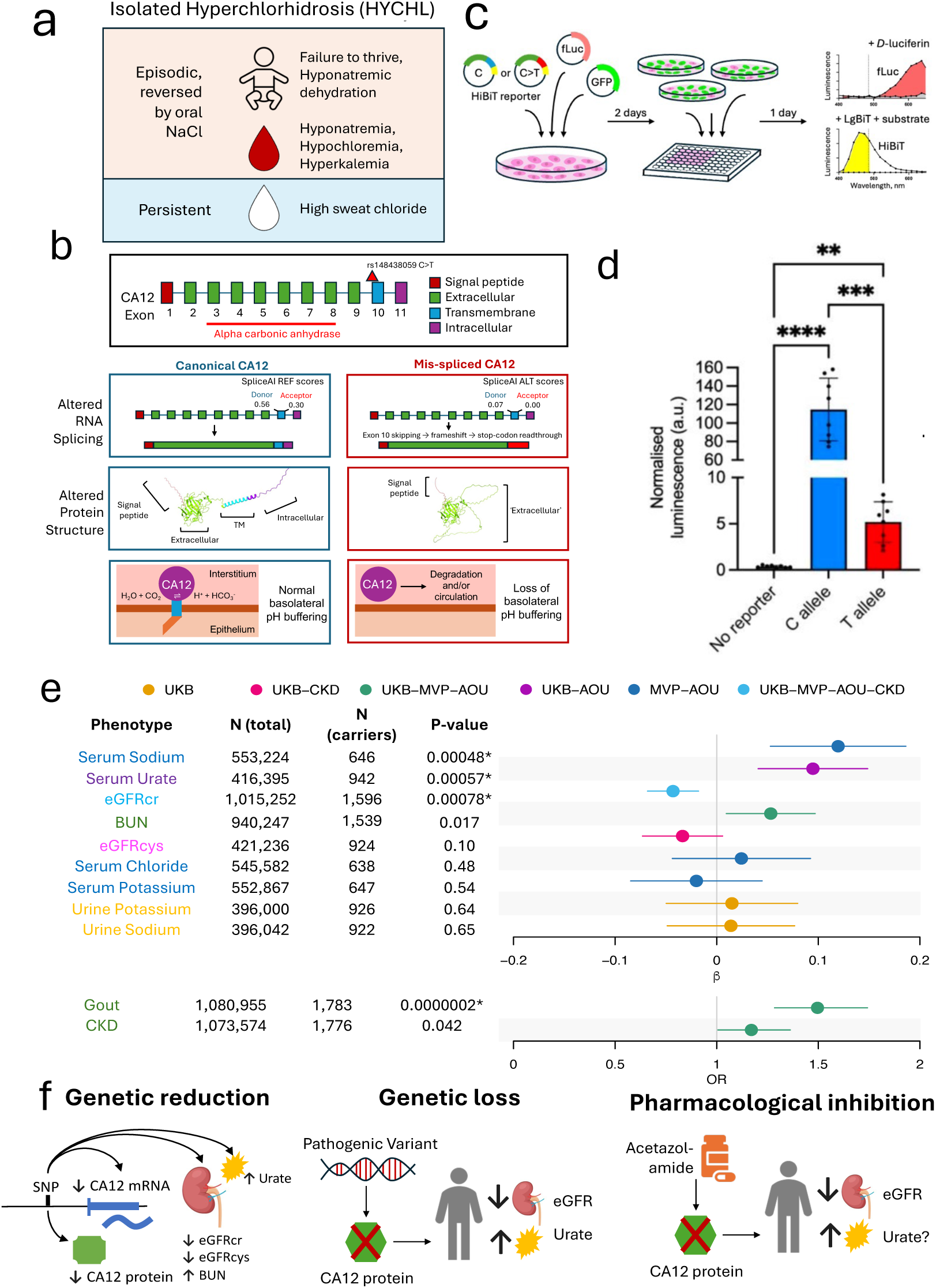
Analysis of a pathogenic CA12 loss-of-function variant recapitulates common CA12 variant associations. **A.** Metabolic symptoms of autosomal recessive isolated hyperchlorhidrosis (HYCHL). **B.** In silico characterization of the pathogenic rs148438059 C>T variant. The rs148438059 variant is shown relative to the CA12 gene structure, along with the effects of the pathogenic T allele on translation of mRNA, protein structure, and protein localization. Protein structures are AlphaFold visualizations of CA12 canonical an abnormal protein (induced by T allele), coloured by domain. **C.** Schematic overview of *in vitro* CA12 variant expression analysis (see Methods for details). **D**. Quantification of proximal tubule cell lysate exogenous CA12 protein level from relative luminescent signal of cells expressing canonical or variant HiBiT-tagged CA12 plasmids. *P* values calculated from one way ANOVA (n = 8 replicates across two experiments) ‘**’ indicates *P* < 0.01, ‘***’ indicates *P* < 0.001. **E.** Phenotypic effects of HYCHL pathogenic variant rs148438059 estimated by meta-analysis of rs148438059 alleles across 4 biobanks (UKB – UK Biobank, CKD – CKDGen, MVP – Million Veterans Program, AOU – All of Us.) Right panel shows estimated beta coefficients (or Odds Ratio – *x* axis) for the associations between of the rs148438059-T allele on metabolic and renal phenotypes (*y* axis). Points, coloured by meta-analysis cohorts, show effect and lines show 95% confidence intervals). ‘*’ indicates calculated *P* values < Bonferroni threshold. **F.** Schematic showing observed effects of CA12 reduction by common genetic variants (left), and a pathogenic loss-of-function variant (middle) on kidney function and urate. Right panel shows hypothesized effects of CA12 inhibition (acetazolamide treatment) on kidney function and urate levels, in line with genetic findings.

As an orthogonal replication analysis, we sought to investigate ‘human knockouts’ who inherited biologically non-functional copies of CA12 alleles at birth. We noted that homozygous inheritance of CA12 loss-of-function variants results in a rare autosomal recessive disorder - Isolated Hyperchlorhidrosis (HYCHL). HYCHL patients present in infancy with failure to thrive, hyponatraemic dehydration, hypochloraemia and hyperkalaemia ^82–85^ (**Fig. 8A**). These symptoms are successfully reversed upon sodium chloride treatment, but patients exhibit a lifelong loss of salt through sweat, owing to the complete ablation of CA activity in sweat glands (where CA12 is the only CA isozyme)^83^. Kidney function in paediatric HYCHL patients was reported as normal, and it was suggested that other CAs expressed in the kidney compensate for renal CA12 loss in HYCHL^86^. However, there is scarce data on health outcomes of HYCHL patients’ past childhood-early adulthood, and the lifelong effects of CA12 loss remain uncharacterized.

We first tracked reported HYCHL alleles in UKBiobank and found that one rare HYCHL allele (rs148438059-T) was present with an allele frequency of 0.001 (1 homozygote and 1038 heterozygotes). We examined phenotypic information for the rs148438059-T homozygous UKBiobank participant, who we found had abnormally low urinary sodium (ranked 2^nd^ lowest of 482,937 UKBB participants), this would be consistent with the loss of salt through sweat, which characterizes HYCHL. We noted an ICD-10 code for haematuria but found no biochemical evidence of reduced eGFR (eGFRcr = 99 ml/min/1.73m2). Urinary albumin was not recorded.

We then sought to examine the effects of the rare allele of rs148438059^84^, on mRNA and protein structure of CA12 *in silico* (**Fig. 8B**). rs148438059-T disrupts the splice acceptor site on exon 10, which encodes the transmembrane domain of CA12. SpliceAI predicted that the rare allele leads to a loss of both splice donor and acceptor sites on exon 10, exon 10 skipping, inducing a frameshift following codon 302 on exon 9. The translation results in readthrough of the native stop codon, until reaching a novel stop codon in the 3’ UTR, adding an extra 111 residues. InterPro^87^ predicted that both canonical and aberrant isoforms have a carbonic anhydrase domain and a signal peptide, but only the canonical isoforms have a transmembrane region. This suggests the novel isoforms resulting from aberrant splicing are mis-localized and do not reside in the transmembrane. Quick2D^88^ predicted that no big differences are expected between the isoform in terms of protein structure but for the lack of the transmembrane helix in the novel isoforms. These results were confirmed with more specific tools: DeepTMHMM^89^, NetSurf-3.0^90^ and CAID^91^. The AlphaFold^92^-predicted structure for the aberrantly spliced CA12 isoform demonstrated loss of the transmembrane domain (**Fig. 8B**), alongside electrostatic and lipophilic properties (**Supplementary Fig. 3**).

The structure of protein product of aberrantly spliced rs148438059 isoforms could render it a potential target for degradation - Lee *et al.* 2016 showed absence of CA12 protein lacking exon 10 in cellular lysates^15^. To test this possibility, we generated reporter plasmids expressing HiBiT-tagged coding sequences corresponding to the common (C allele) and aberrantly spliced (T allele) CA12 isoforms and compared their relative protein expression level in a proximal tubule cell line (**Fig. 8C, Supplementary Fig. 4**). Exogenous expression of the C allele reporter plasmid resulted in a strong luminescent signal in cell lysate, whilst luminescence levels were dramatically lower, but detectable, in cell lysate from cells expressing the T allele reporter plasmid (**Fig. 8D**), suggesting reduced protein stability and/or targeted degradation. This supports the pathogenic role of rs148438059 HYCHL allele as the cause of significant loss of CA12 on the basolateral membrane of renal tubule cells.

To further investigate phenotypic consequence of HYCHL carrier status, we further tracked rs148438059 in All of Us^93^ (AoU), Million Veterans Program^94^ (MVP) and CKDGen^95^, and identified further carriers of European ancestry in these cohorts (**Fig. 8E**). In each cohort we examined the association of rs148438059-T with available HYCHL-relevant electrolytes alongside CKD-defining traits and ICD-10 diagnoses (**Fig. 8E**). Our meta-analysis revealed that carriers of rs148438059-T had significantly decreased eGFRcr (*P* = 7.8×10^-4^) and significantly increased serum urate (*P* = 5.7×10^-4^), together with 1.5-fold increased odds of gout relative to non-carriers (*P* = 4.5×10^-5^). Intriguingly, HYCHL carriers had significantly increased serum sodium levels (*P* = 2.2×10^-7^), which could indicate a compensation from CA12-associated salt loss e.g. by dietary sodium intake. We found further nominally significant and biologically consistent associations with decreased BUN (*P* = 0.017) and increased odds of CKD (*P* = 0.042). Altogether, these data suggest that: (i) HYCHL alleles are not health-neutral in a heterozygous conformation, (ii) HYCHL carriers may display a mitigated disease phenotype, (iii) metabolic and renal abnormalities may form part of the phenotypic spectrum of HYCHL in adulthood, and (iv) these phenotypic consequences may arise due to the reduction of CA12 expression in the kidney.

Our results from integrative multi-omics, human genetics and *in silico* functional analysis provide an orthogonal support for a model where genetic reduction of CA12 in the human kidney leads to decreased renal function and increased circulating urate (**Fig. 8F**). This is partially supported by clinical observations; acetazolamide (inhibitor of carbonic anhydrases including CA12) may lead to a drop in GFR ^96,97^. The effect of acetazolamide on urate is less clear: one study suggested mild uricosuric effects owing to urinary alkalization ^98^, whilst others reported hyperuricemia and gout as a consequence of acetazolamide treatment^99,100^. Serum urate elevation is a common adverse effect of thiazide and loop diuretics^101^. Altogether, our findings reveal a novel role for CA12 as a key regulator of renal function and urate handling. Furthermore, indirectly they suggest that pharmacological CA12 inhibition by acetazolamide may indeed have potential for adverse effects of relevance to patients at risk of hyperuricemia and gout (**Fig. 8F**).

## Discussion

In this study, we traced the effects of genetic variants through the regulatory cascade of DNA methylation, mRNA expression and protein abundance, to prioritize genes which drive the genetic predisposition to CKD. We followed a multi-level strategy embedded in the convergence of four traits which define kidney health and disease in clinical practice.

We reasoned that by triangulating genetic associations from multiple CKD-defining traits, we would capture the genetic variation which best proxies the endophenotype of CKD. Indeed, we found that genomic regions associated with multiple CKD-defining traits preferentially harbour CKD kidney genes, compared to regions linked only to single CKD-defining traits. Furthermore, genes prioritized by more than one CKD-defining trait showed the greatest: (i) specificity to the kidney, (ii) functional constraint, (iii) enrichment for function in renal development, and (iv) overlap with single gene causes of renal disease. In contrast, genes which were specific to a single trait (most commonly eGFRcr), showed numerically weaker enrichment for all above characteristics.

Our approach to GWAS of CKD is perhaps more stringent than several other eGFRcr-focused studies of kidney disease^5,11,12^, which typically define support for eGFRcr by nominally significant (*P* < 0.05) and directionally consistent association with additional trait(s) (BUN, eGFRcys). One may argue that being overly permissive towards variants primarily associated with creatinine metabolism, may result in under-reporting of signals of CKD-susceptibility which are not driven by creatinine. Notably, our study found that ∼38% of genes prioritized for eGFRcr lacked support from another CKD-defining trait. Furthermore, these eGFRcr-specific genes, when examined in the context of DNA methylation, were over-represented for function in muscle development, which can reflect (at least to some extent) the relationship between muscle mass and creatinine levels. Interestingly, we also prioritized 14 kidney genes with support from CKD-defining traits not based on creatinine.

Our studies provide mechanistic insight into how GWAS variants transmit their effects through the epigenome, transcriptome and proteome to CKD. A controlled analysis of heritability mediation revealed that *cis*-mQTL, *cis*-eQTL and *cis*-pQTL all mediate similar portions of heritability after accounting for the analysis’ sample size and number of included features (CpGs, genes, proteins). One possible interpretation for this result could be that that the regulatory effects embedded in each molecular layer may be largely overlapping. However, it is important to distinguish between the technology-driven disparity in the number of genes and proteins in transcriptomics and proteomics, and the biologically meaningful disparity in the number of CpGs and genes. The methylation of additional CpG sites, outside of gene promoters, captures information on gene regulation driven by enhancer and silencer elements, including those specific to developmental timepoints and environments. For this reason, the mediated heritability estimated from the ∼350k measured CpGs – and particularly that estimated from the set of ∼5k promoter CpGs – are underestimates of the true heritability mediated by DNA methylation (due to the incomplete coverage of DNA methylation arrays). Consistent with our prior analysis of blood pressure, we find that little CKD GWAS heritability is mediated by microRNAs^17^. Furthermore, whilst all estimates of molQTL-mediated heritability are low in absolute terms, these are likely underestimated due to a downward bias from the sample size. We suspect that the remaining ‘unmediated’ heritability may be due to the effects of trans molQTL, alongside cell-types and context-specific molQTL. Expanding molQTL datasets (in both the depth and breadth) to capture these effects should be the future focus of the field.

Our integrative analyses spanning multiple layers of kidney omics permitted us to make several important observations which would have not been possible from single omics alone. Most notably, a proportion of genetic variants which predispose individuals to CKD act by influencing kidney DNA methylation, but not gene expression in adult tissue. These genes show a strong enrichment for function during kidney development, which suggests that they may operate by modulating expression at the embryonic or fetal stages, and that transcriptome profiling of adult human kidney will not be able to capture such effects. These observations support that kidney DNA methylation patterns retain genetic memory of embryonic gene regulation. This highlights a need to develop resources and strategies to capture the effects of DNA methylation, with potential importance to human health and disease beyond CKD^68,69^.

Finally, we illustrate how findings from analyses of common variants, when combined with omics and large datasets, can: uncover unknown clinical dimensions of monogenic forms of human disease (e.g. increased risk of gout and CKD in heterozygote carriers of a mutant allele for isolated hyperchlorhidrosis), illustrate biologically consistent effects of both extremely rare and common allelic variants of the same gene (e.g., *CA12*) on CKD; and provide insights of potential therapeutic importance to CKD and coexistent diseases (e.g. a potential increase in risk of hyperuricaemia associated with treatment with acetazolamide).

We are aware of several limitations of our study, including its use of European-ancestry participants only. This was necessary to match the GWAS population to our collection of kidney samples which are overwhelmingly from European-ancestry donors. To extend our findings into other ancestry groups, future efforts will focus on multi-ancestry GWAS and molQTL mapping – these require the collection of kidney tissue from under-represented ancestries^102^. Further work will also aim to separate multiple signals within GWAS and molQTL locus regions, to increase the yield of GWAS target genes discovered by colocalization. We were also limited by the relatively small size of the proteomics dataset, and its limited overlap with other molecular layers (particularly methylation). The small sample size meant that our proteomics data were underpowered to detect many pQTLs and assess their colocalizations with GWAS and kidney eQTLs. However, our findings here for CKD, and elsewhere for blood pressure^17^, show that many genes nominated by kidney mRNA are supported at the level of kidney protein – which highlights the therapeutic potential of targets. A goal of future studies will be to expand kidney tissue proteomics, to establish the relationship between the renal methylome, transcriptome and proteome, and characterize kidney proteins as the ultimate effectors of the causal molecular chain.

## Methods

### Populations and phenotypes - UKBB

The eGFRcr for UKBB participants was calculated using the CKD-EPI creatinine equation in R using the ‘CKDEpi.creat’ function from the ‘nephro’ R package^103^, after converting serum creatinine from umol/l to mg/dl. This applied the 2009 CKD-EPI equation, which has been shown to produce less-biased estimates of glomerular filtration, compared to eGFRcr equations without race^104,105^. eGFRcr was winsorized to 15 and 200 ml/min/1.37m^2^. The rank-based inverse normal transformed residuals of the natural logarithm of eGFRcr adjusted for age and sex was used as the continuous trait for the GWAS^3^. Using cystatin C (UKBB field ID: 30720, mg/L), eGFRcys was calculated using the CKD-EPI cystatin C equation available as R command ‘CKDEpi.cys’ in ‘nephro’ package^104^. Residuals of the natural logarithm of eGFRcys adjusted for age and sex were used as the continuous trait for the GWAS^106^. UKBB BUN concentrations (field ID: 30670, mmol/L) were converted into mg/dL by dividing by 0.3571. The rank-based inverse normal transformed residuals of log-transformed BUN adjusted for age and sex were used as the dependent trait in GWAS^3^. UKBB urinary concentrations of albumin and creatinine were derived from field ID: 30500 (mg/L) and field ID: 30510 (µmol/L), respectively. The missing values were imputed using the minimum detected value for the corresponding trait. The urinary albumin-creatinine ratio (UACR) was expressed in mg/g and calculated as urinary albumin (mg/L) / urinary creatinine (mg/dL = µmol/L x 0.0113) x 100, in line with the previous study^4^. The rank-based inverse normal transformed residuals of the natural logarithm of UACR adjusted for age and sex were used as the continuous trait for the GWAS^4^.

### Genome-wide association analysis in UKBB

Linear mixed models (i.e., BOLT-LMM^107^) were employed to conduct the GWAS, regressing the transformed phenotype against the imputed genetic variant. We used UKBB version 3 imputed genotype data, which used a reference panel combining HRC, UK10K and 1000 Genomes references. An additive model of inheritance was specified, testing variants with MAF > 0.5% and INFO score > 0.3, and adjusting for covariates including age, sex, genotyping array kit, and the first ten genetic principal components. Genetic ancestry was inferred using the ‘inference’ function in KING v.2.2.4, and defined based on probability ≥0.95^108^. Individuals in UKBB were excluded from our analyses based on the following criteria^109^: (1) outliers in heterozygosity (field ID: 22027); (2) individual calling rate (calculated using probe sets internal to Affymetrix) < 97% or the resolution of the distributions of intensity ‘contrast’ value < 0.82; (3) carriers of sex chromosomal abnormalities (i.e., configurations other than XX or XY); (4) within 3^rd^ degree of relatedness with other individuals; (5) non-European genetic ancestry.

### CKDGen GWAS summary statistics

For each kidney marker, the GWAS summary statistics from the CKDGen consortium round 4 analyses were downloaded from the CKDGen data portal (https://ckdgen.imbi.uni-freiburg.de). Details on the phenotype preparation and covariate adjustment are detailed in the original studies^3–5^.

### Meta-analyses

The GWAS significance levels for UKBB and CKDGen were corrected using the LD-score regression intercept from LDSC^110^, to mitigate potential population stratification bias. LD-score regression was fitted based on European ancestry participants from the 1000 Genomes project^111^, as a reference, using publicly available (https://zenodo.org/records/10515792) precomputed LD scores. Fixed-effect inverse-variance weighted meta-analyses was performed using METAL^112^, combining the GWASs of each corresponding phenotype from UKBB and CKDGen. The meta-analysed GWAS results were not further corrected with the LD-score intercepts. We retained variants with heterogeneity I^2^ < 75% in the meta-analysed results for the downstream analyses.

### Locus definition and identification of non-overlapping locus regions

In each GWAS meta-analysis result, variants exceeding the genome-wide significance threshold (*P*=5×10^-8^) were collected and listed in ascending order in each chromosome. To define independent loci, genetic variants were first assigned to genomic windows, which were defined by the genetic coordinate (in genome build hg19) +/- 500Kb. For each chromosome, iteratively, variants with overlapping windows were merged into one locus – defined by the minimum lower boundary and maximum upper boundary. Locus boundaries for each GWAS were merged using bedtools^113^ to identify non-overlapping CKD locus regions. Each non-overlapping locus region was annotated by phenotype based on at least one variant with *P*=5×10^-8^ within its boundaries. Both overlapping and unique locus regions were visualised using UpsetR^114^.

### Estimation of SNP-based heritability, global genetic correlations, and heritability enrichment

LDSC was used to estimate the SNP-based heritability, global genetic correlations, and partitioned heritability of each phenotype. LDSC ‘munge_sumstats.py’ was first applied to GWAS summary statistics to calculate z-scores and filter for HapMap3 SNPs (downloaded from https://zenodo.org/records/10515792). LD-score regression was then performed to estimate heritability and genetic correlations using the European-Ancestry 1000 Genomes reference of all EUR samples and precomputed LD scores^24,110^. To assess the enrichment of heritability in tissue-specific gene expression, we performed stratified LD-score regression applied to specifically-expressed genes (LDSC-SEG^115^) using cell-type/tissue annotations downloadable at https://console.cloud.google.com/storage/browser/broad-alkesgroup-public-requester-pays.

### Estimation of local bivariate genetic correlations

Local genetic correlations between phenotype-pairs were computed using LAVA^25^. The input to this analysis included the HapMap3 filtered GWAS summary statistics generated during LDSC analysis, the 1000 Genomes European ancestry reference, used for LDSC analysis, the 508 non-overlapping locus regions, and a matrix of estimated sample correlations, also calculated in the previous LDSC analysis. LAVA univariate tests were first performed to identify locus-trait-pair combinations with sufficient genetic signal for both traits within a region. LAVA bivariate tests were then performed for 833 locus region-trait-pair combinations (with Bonferroni-adjusted *P* < 0.05) to estimate local genetic correlations.

### Human Kidney Tissue Resource – populations, samples and ethical compliance

The Human Kidney Tissue Resource (HKTR) serves as a repository of human kidney tissue samples curated for multi-omics analyses^6–8,26,102,116–122^. The resource is based on contributions from the following studies: the TRANScriptome of renaL humAn TissuE study (TRANSLATE^17,116,117,123^), TRANScriptome of renaL humAn TissuE - Transplant study (TRANSLATE-T^8,118^), moleculAr analysis of human kiDney-Manchester renal tIssue pRojEct (ADMIRE^118^), Renal gEne expreSsion and PredispOsition to cardiovascular and kidNey Disease (RESPOND^118^) and moleculaR analysis of mEchanisms regulating gene exPression in post-ischAemic Injury to Renal allograft (REPAIR^118^). The specimens used in this study were sourced either from the healthy (non-cancerous) pole of the kidney, obtained immediately following elective nephrectomy, or from needle biopsies of donor kidneys before transplantation. There were 478 specimens in total including 372 from nephrectomies and 106 from kidney biopsies. All participants were of European ancestry. Further details on the individuals enrolled in each study are available in **Supplementary Table 9**.

### National Institutes of Health kidney collections – populations, samples and ethical compliance

This study used data from normal adjacent tissue (NAT) kidney samples, which were collected by from renal cancer patients after nephrectomies. Data from kidney NAT samples were generated by The Cancer Genome Atlas (TCGA^27^) and Clinical Proteomics Tumor Analysis Consortium (CPTAC^28,124^), which were accessed under the approved dbGAP project 13040. Data relating to TCGA and CPTAC participants were obtained from the National Cancer Institute Genomic Data Commons (GDC^125^) and Proteomic Data Commons (PDC^126^). Patient characteristics relating to 91 TCGA and 83 CPTAC samples are described in **Supplementary Table 9.**

### DNA extraction, genotyping and whole genome sequencing, processing and quality control

HKTR kidney tissue samples were homogenized and subject to DNA extraction using the Qiagen DNeasy Blood and Tissue Kit and subsequently hybridised to the Illumina HumanCoreExome-24 BeadChip array, as reported before^17,26^. Genotype calls were made using Illumina GenomeStudio. DNA quality control included excluding samples with cryptic relatedness, a genotyping rate below 95%, heterozygosity outside ±3 standard deviations from the mean, or a mismatch between genetic and phenotypic sex, as per our previous studies^17,26^. Variants with genotyping rate < 95%, genomic locations on a sex chromosome or the mitochondrial genome, ambiguous genomic positions, Hardy Weinberg Equilibrium (HWE) *P* values < 1 × 10^-3^, and MAF < 5% were excluded from the variant-level quality control. Genotype imputation was conducted on Michigan Imputation Server (MIS^127^) using the 1000 Genomes phase 3 reference panel^111^. Variants with duplicated genomic location, imputation score <0.4, MAF < 1% or HWE *P* value < 1 × 10^-6^ were excluded from the post-imputation quality control. After all genotyping quality control processes, 8,735,852 variants were retained. EIGENSTRAT^128^ and SNPWeights^129^ were used to derive the genotype principal components from genotyped autosomal variants that passed all genotyping quality control filters.

In TCGA, DNA was extracted from blood samples using QiAamp Blood Midi Kit and hybridised with probes on the Affymetrix SNP 6.0 array (composed of 906,600 probes); genotype calls were made using the Birdseed algorithm^130^. The TCGA genotype data were downloaded from the GDC Portal’s legacy archive. A total of 525 files were initially identified using the following query criteria: (1) project name: TCGA, (2) primary site: kidney, (3) race: white, (4) sample type: solid tissue normal, (5) experimental strategy: genotyping array and (6) access: controlled. Genotype data were initially downloaded for 110 NAT samples with matching RNA-sequencing data. Genotype quality control, imputation, post-imputation quality control and genotype principal component analyses were performed in line with the protocol used in the HKTR. After all steps were complete 8,541,201 variants remained in the TCGA genotyping dataset.

DNA extraction and whole genome sequencing (WGS) in CPTAC was performed on blood samples for each sample according to the CPTAC standard operating procedures (available at https://brd.nci.nih.gov/brd/sop-compendium/show/41). Briefly, DNA was extracted using the QIAsymphony DNA Mini Kit (Qiagen), acoustically sheared, indexed, multiplexed and then sequenced to 15x coverage on an Illumina HiSeqX. FASTQ reads were mapped to the National Cancer Institute (NCI) GDC GRCh38.d1.vd1 reference sequence (https://api.gdc.cancer.gov/data/254f697d-310d-4d7d-a27b-27fbf767a834) following the standard GDC protocol (https://docs.gdc.cancer.gov/Data/Bioinformatics_Pipelines/DNA_Seq_Variant_Calling_Pipeline/#alignment-workflow) which involves read mapping with BWA-MEM^131^, alignment sorting and merging using Picard (https://broadinstitute.github.io/picard), duplicate marking (also using Picard) and finally base quality score recalibration using the Genome Analysis ToolKit (GATK^132^).

WGS BAM files generated through this workflow were then downloaded from the GDC portal using the following query: (1) project name: CPTAC, (2) primary site: kidney, (3) sample type: solid tissue normal, (4) experimental strategy: WGS, (5) race: white and (6) data format: BAM. Variant calling was performed on these files with HaplotypeCaller from GATK^132^ v3.8 in single sample GVCF mode with the arguments: “-dontUseSoftClippedBases -ERC GVCF”. All individual GVCF files were then genotyped jointly using the GATK GenotypeGVCFs tool with default arguments. BCFTools^133^ was then used to split multiallelic variant calls into biallelic calls, producing 19,847,739 variants in the CPTAC dataset.

### RNA-sequencing data generation, processing, normalization and quality control

RNA was extracted from HKTR kidney tissue samples using either Qiagen RNeasy or mirRNeasy kits. 1µg of RNA was used as input to either the NEBNext Poly(A) mRNA isolation kit (New England Biolabs) or the Illumina TruSeq RNA Library Prep RNA kit (Illumina). Libraries were then sequenced on Illumina instruments using either 100bp paired-end reads (Illumina HiSeq 2000), 75bp paired-end reads (Illumina NextSeq) or 150bp paired-end reads (Illumina NovaSeq), as reported before^17,26^.

For all TCGA samples, RNA was extracted from snap-frozen tissue samples following the Biospecimen Research Database Standard Operating Procedure (SOP) “M001” for “DNA/RNA Extraction with Allprep (DNA) and MirVana (Total RNA with Small RNA)”. Poly-A selected sequencing libraries were sequenced on an Illumina HiSeq2000 providing an average of 81 million paired reads per sample. Aligned RNA-sequencing reads were obtained from TCGA via the GDC data portal using the query: (1) project name: TCGA, (2) primary site: kidney, (3) experimental strategy: RNA-Seq, (4) sample type: solid tissue normal, (5) race: white, (6): data category = raw sequencing data, (7) data type: aligned reads.

RNA was extracted from CPTAC samples following the study’s standard operating procedures (https://brd.nci.nih.gov/brd/sop-compendium/show/41). Libraries were prepared using the Illumina TruSeq Total RNA protocol with rRNA depletion (RiboZero Gold) and sequenced on an Illumina HiSeq 4000, generating at least 120 million paired-end 75bp reads per samples. Aligned CPTAC RNA-seq data were obtained from the GDC data portal using the query: (1) project name: CPTAC, (2) primary site: kidney, (3) experimental strategy: RNA-Seq, (4) sample type: solid tissue normal, (5) race: white, (6) data category = sequencing data, and (7) data type: aligned reads.

FASTQ reads from TCGA and CPTAC samples were extracted BAM files using the biobambam2^134^ command “bamtofastq”. HKTR, TCGA, and CPTAC FASTQ reads were pseudoaligned to the GRCh38 genome and Ensembl release 83 transcriptome references using Kallisto^135^. Gene expression was first quantified in units of transcripts per million (TPM) at the transcript level. In line with our previous studies, gene-level expression values were calculated as the sum of TPM values for all transcripts, per gene^17,26,118,122^. TPM values were then log_2_ transformed after the addition of a constant value of 1.0, to avoid log transformation of zero values. Genes were annotated with chromosome positions and biotypes from Ensembl 111 GRCh37 build and filtered for: 1) log_2_(TPM+1) > 0.1 and read count ≥ 6 in at least 20% of HKTR, TCGA and CPTAC samples, 2) autosomal position, and 3) protein-coding and lncRNA biotype. A total of 19,428 genes passing these criteria were retained for further analysis.

We applied four different quality control processes to the log-transformed TPM values to ensure sample quality and identity. Samples were selected for: 1) ≥ 10 million pseudoaligned paired reads, 2) D-statistic (measuring intra-batch transcriptomic variability^136^) ≤ 5, 3), consistent expression of sex specific genes (females: XIST expression, males: expression of MSY genes *RPS4Y1*, *DDX3Y*, *EIF1AY*, KDM5D, *USP9Y*), and 5) consistent genotype calls from DNA (see above) and RNA (called using the GATK^132^ RNA-seq variant calling pipeline). A total of 478 HKTR, 91 TCGA, and 76 CPTAC samples passed sample-level RNA-seq quality control.

### DNA methylation data generation, processing, normalization and quality control

For HKTR kidneys, 750ng of high-quality DNA was extracted and processed using the Zymo EZ DNA Methylation Kit (Zymo Research) for bisulphite treatment in accordance with manufacturer guidelines relating to alternative incubation conditions for Illumina methylation arrays, as reported before^26^. Bisulphite-converted DNA was hybridised using Infinium HumanMethylation450 BeadChip (Illumina; 96 sample kit – 96 samples) and MethylationEPIC BeadChip (Illumina; 96 sample kit – 284 samples) arrays and imaged using the high-throughput iScan System (Illumina).

Methylation profiles for TCGA NAT kidneys were retrieved from the GDC data portal with the following criteria: (1) project name: TCGA (2) primary site: kidney (3) sample type: solid tissue normal (4) race: white (5) ethnicity: not Hispanic or Latino (6) data category: raw microarray data (7) data type: raw intensities (8) experimental strategy: methylation array (9) data format: idat (10) platform: Illumina Human Methylation 450.

Raw methylation data for HKTR and TCGA samples were merged using minfi^137^ and quantified into M-values, which were then normalized with the ‘dasen’ method from the R package ‘wateRmelon’ ^138^. Starting from 452,567 CpG probes shared between the two methylation arrays, probes were selected for downstream analysis based on: (1) detection *P* value < 1×10^-16^, (2) autosomal CpG position, (3) no cross-reactivity with multiple genomic regions, and (4) no overlap with common genetic variants (MAF > 10%). After application of these filters, 370,711 CpG probes were available for further testing.

Samples were included for further analysis based on: (1) consistent sample labelling, (2) matching genotype information post-quality control, (3) methylation rate ≥ 95%, and (4) CpG probe detection rate ≥ 5% (determined by detection *P* value < 1×10^-16^). A total of 350 HKTR and 16 TCGA methylation samples survived quality control.

### Proteomics data generation, processing, normalization and quality control

The processed CPTAC proteomics data were obtained from the supplementary material of Li et al. 2024, with all data being generated and processed as reported in full in the original publication^139^.

Protein abundance matrices were filtered for kidney NAT samples with matching genotype data which were previously identified using the GDC/PDC query: (1) project name: CPTAC, (2) primary site: kidney, (3) sample type: solid tissue normal, (4) experimental strategy: WGS, (5) race: white and (6) data format: BAM. A total of 83 samples were included in downstream analysis. 8,362 detected proteins were filtered for missingness < 20%, resulting in a total of 5,990 proteins included in downstream analysis.

### miRNA sequencing data generation, processing, normalization and quality control

miRNA sequencing libraries were prepared for HKTR RNA samples with Illumina TruSeq and sequenced with 75bp single-end reads on Illumina NextSeq, or 50bp single-end reads on Illumina HiSeq2500, as reported before^17^.

TCGA microRNA sequencing libraries were prepared from extracted RNA and sequenced using 30bp single-end reads on an Illumina HiSeq2500 platform. Aligned miRNA sequencing reads from TCGA via the GDC portal using the query: (1) project name: TCGA, (2) primary site: kidney, (3) race: white, (4) sample type: solid tissue normal, (5) experimental strategy: miRNA-seq, and (6) data format: BAM. FASTQ reads were extracted BAM files using the biobambam2^134^ command “bamtofastq”.

Trimmomatic^140^ was first used to remove standard Illumina adapter sequences from HKTR and TCGA FASTQ files. FASTQ reads were mapped to the GRCh38 human genome reference sequence using STAR v2.5.3a ^141^. STAR v2.5.3a was also used to quantify mature miRNAs from miRbase^142^ release 22. Reads per million values (RPM) were calculated from the STAR read counts and then log_2_ transformed after addition of a constant value of 1 RPM. log_2_(RPM+1) values from HKTR and TCGA were corrected for batch effect using the ComBat^143^ function from the ‘sva’ R-package. Mature miRNA transcripts were filtered for: (1) log_2_(RPM+1) > 0.1 in greater than 10% of HKTR or TCGA samples, (2) autosomal position, and (3) non-zero expression inter-quartile range. A total of 1,449 mature miRNAs were retained for downstream analysis.

Sample-level quality control was performed by selecting samples with: (1) ≥ 2 million aligned reads, and (2) D-statistic^136^ ≤ 5. A total of 327 HKTR and 82 TCGA samples passed the quality control.

### Cis-Expression Quantitative Trait Loci Analysis

A total of 645 samples with matched genotype and RNA-seq data were identified from HKTR, TCGA and CPTAC. To obtain a common set of variants for *cis-*eQTL analysis, 478 HKTR, 91 TCGA, and 76 CPTAC genotype datasets were merged with the BCFtools^133^ “merge” command, and filtered with PLINK2^144^ for variants with: (i) MAF > 1%, (ii) HWE *P* > 0.001, and (iii) genotype call rate > 95%. Gene expression levels in corresponding samples were further normalized by robust quantile-normalization (using medians) of log2 TPM values with aroma.light^145^, followed by RBINT.

To discover *cis*-eQTLs, we tested autosomal variants for association with normalized expression of nearby genes (defined by TSS within 1Mb), adjusting for age, sex, study and 3 genetic principal components (calculated from merged genotypes with KING^108^). To control for potential unobserved confounders, such as uncorrected batch effects, c*is*-eQTL associations were adjusted for the gene expression principal components:^146^ we first calculated gene expression principal components from normalized RNA-seq data, and then varied the number of PCs included in eQTL regression models from 0 to 120 (in increments of 10). This found that the number of significant eGenes (*FDR* Q < 0.05) did not significantly increase past inclusion of 80 PCs in eQTL analysis (i.e. the ‘elbow point’)^146^. Our *cis-*eQTL analysis was performed using QTLtools^147^ v1.3. This used a ‘permutation analysis’ to identify the number of significant eGenes with the command “permute 1000 -normal -window 1000000’. To correct for the number of genes tested, beta-approximated *P* values were corrected by Storey’s method^148^. *cis*-eQTLs were then uncovered using the command ‘nominal -normal -window 1000000’. To define significant eQTLs, we defined nominal *P* value thresholds for each gene by performing Benjamini-Hochberg (BH) correction^149^ of beta-adjusted *P* values at *FDR* < 0.05.

### Cis-Protein Quantitative Trait Loci – data curation, normalization and QTL mapping

PLINK2^144^ was used to filter variants in 83 CPTAC genotypes for: (1) MAF > 10%, (2) HWE *P* > 0.001, and (3) genotyping rate > 95%. Log_2_ protein abundances were annotated with chromosome positions and gene identifier from Ensembl 111 GRCh37 build. Our *cis*-pQTL analysis was performed using QTLtools as described for *cis*-eQTL analysis to discover variants in association with the abundance of kidney proteins (pGenes) encoded by a nearby gene (TSS <1Mb). The correction for unknown confounders was optimized by varying the number of protein PCs included in the model from 0-20, in increments of 1. Protein PCs were calculated from all proteins with no missing values. We found that pGene discovery decreased after inclusion of 12 PCs. Hence, we included 12 PCs in the final *cis*-pQTL model. All QTLtools parameters and thresholds were the same as for *cis*-eQTL analysis.

### Cis-Methylation Quantitative Trait Loci Analysis

366 samples with matched genotype and CpG methylation data were identified from HKTR and TCGA. Genotypes were merged and filtered, as described for eQTL analysis, using the following thresholds: (1) MAF > 5%, (2) HWE *P* > 0.001, and (3) genotyping rate > 95%. Methylation M-values in corresponding samples were further normalized by RBINT and CpG positions were obtained from the HM450 array annotation file. *Our cis*-mQTL analysis was performed using QTLtools, as described for *cis*-eQTL analysis (with the same parameters and thresholds), to discover autosomal variants associated with the methylation of nearby (<1Mb) CpG sites (mGenes). We optimised correction for unknown confounders by including variable numbers of methylation PCs in the mQTL model (0-50, incrementing by 5). A total of 30 methylation PCs were included in the final model since mCpG discovery decreased after inclusion of 30 PCs.

### *Cis*-miRNA Quantitative Trait Loci – data curation, normalization and QTL mapping

409 samples with matched genotype and miRNA-seq data were identified from HKTR and TCGA. Genotypes were merged and filtered, as described for eQTL analysis, using the following thresholds: (1) MAF > 5%, (2) HWE *P* > 0.001, and (3) genotyping rate > 95%. Mature miRNA expression values in log_2_(RPM+1) were obtained for corresponding samples, and subject to further normalization by robust quantile normalization and RBINT. miRNA gene positions were obtained from miRbase release 22^142^. *Our cis*-mirQTL analysis was performed using QTLtools as described above (with the same parameters and thresholds as in the other molQTL analyses). We tested autosomal variants for association with the expression of nearby kidney miRNAs (miRNA gene start <1Mb). We varied the inclusion of miRNA PCs in the final model from (0-10, incrementing by 1) and selected an optimum number of 4 miRNA PCs for inclusion in the final model.

### Estimation of mediated heritability

Heritability mediation analysis was performed for each GWAS of CKD-defining trait using *cis*-molQTL associations by mediated expression score regression (MESC^29^). As the input, molecular phenotypes (CpG methylation, gene expression, protein abundance miRNA expression) were normalized as described for molQTL analysis, and adjusted for the same set of covariates as in the final molQTL model (age + sex + study + 3 genetic PCs + *X* molecular PCs) using the “QTLtools correct” command^147^. Genotype PLINK files (for each molQTL analysis) were filtered to only include SNPs present in all analyses. MESC was first used to estimate expression scores (here generalised to ‘molecular scores’) from the genotypes and corrected molecular phenotypes, using genotypes from European ancestry 1000 genomes^111^ participants as a reference. MESC was then applied to estimate the proportion of GWAS heritability mediated by molQTL associations from molecular scores and GWAS z-scores (generated and filtered for HapMap3^150^ SNPs during LDSC analysis). MESC (with the default parameters) was first applied to each molQTL using the full sample size, then (for a balanced comparison), MESC was applied to eQTL, mQTL and mirQTL datasets using a randomly down-sampled set of 83 kidneys (matching the smallest pQTL dataset). Additional sensitivity analyses were performed using down-sampled feature sets and sample sizes: eQTL, pQTL and mQTL datasets were matched for 5,653 common genes (or promoter regions); the mQTL dataset was randomly down-sampled for CpGs (from 50-350 in increments of 50); and the eQTL dataset was randomly down-sampled for sample size (from 50-650, in increments of 50).

### Colocalization

Colocalization was performed between GWAS associations (BUN, eGFRcr, eGFRcys, UACR) and overlapping kidney quantitative trait loci (mQTL, eQTL, and pQTL), intersecting the 508 independent locus regions with *cis* windows for all molecular phenotypes with significant molQTL effects (*Q* < 0.05). We then assessed colocalization within these windows between significant molecular phenotypes and all four GWAS phenotypes, using the Bayesian “coloc.abf” function with default prior values and parameters from the coloc v5.2.3 R package^151^. Evidence of colocalization was defined by posterior probability of colocalization (i.e. PPH4) > 0.8.

### Characterization of CKD-eGenes

To functionally annotate kidney CKD-eGenes, we obtained gene IDs for 406 genes with ‘kidney-elevated’ expression as defined by the Human Protein Atlas. We then tested the enrichment of these kidney-elevated genes by two-sided Fisher’s exact test for 59 multi-trait CKD-eGenes and 189 trait-specific CKD-eGenes compared to 5,733 kidney ‘non-colocalizing’ eGenes (defined by PP H4 > 0.8 for all traits). LOUEF scores of intolerance to loss-of-function variation were obtained from gnomad v4.1^31^ and defined relative to the MANE select transcript^152^. LOEUF scores for multi-trait eGenes (and trait-specific eGenes) were compared to those for non-colocalizing eGenes by a two-sided Mann-Whitney U test.

Single cell expression data were obtained from the integrated single nucleus and single cell RNA-seq atlas of adult kidney tissue from Lake et al.^153^ and downloaded from the cellxgene portal^154^. Seurat^155^ was used to obtain log normalized ‘pseudobulk’ expression values for 52 protein coding multi-trait CKD-eGenes in 16 pre-defined cell types, aggregating expression from healthy, injured, and diseased kidney cells.

### Integration of CPTAC kidney proteomes and transcriptomes

To investigate the extent of kidney transcriptomic and proteomic concordance, normalized gene expression profiles from RNA-seq of renal CPTAC samples were obtained from the supplementary material of Li et. al 2024^139^. The RNA-seq matrix was log_2_ transformed and filtered for 76 NAT samples with matching proteomics data (used in pQTL analysis) and 5,973 genes with matched protein measurements. We used Pearson’s test to calculate the correlation between median log_2_ normalized protein abundance and median log_2_ normalized mRNA abundance. In addition, 5,973 individual Pearson’s correlations were calculated for each kidney mRNA-protein pairs; the generated *P* values were subject to BH-correction^149^ (*FDR* < 0.05).

### Characterization of kidney cis protein quantitative trait loci

The intra-pQTL distance (from pSNP to the target gene’s TSS) was compared to the respective intra-eQTL distance, using the kidney eQTL data from 645 kidney samples by a two-sided Mann-Whitney U test. This test was also used to compare the number of pGenes/eGenes partnered to each lead pSNP/eSNP (respectively).

To functionally annotate kidney pSNPs we used variant consequence, genomic context, epigenomic marks and chromatin states in Ensembl Variant Effect Predictor (VEP)^156^ build 110 GRCh37. The Ensembl VEP command line tool was used to annotate variants with consequences (e.g. missense, synonymous) and genomic contexts (e.g. intronic, UTR) relative to the Matched Annotation from NCBI and EMBL-EBI (MANE) select transcript for the nearest protein coding gene^152^. Variants were supplied to VEP in VCF format, and to provide kidney-specific epigenomic annotations. VEP was configured with BED files (VEP option ‘--custom’) obtained from ENCODE^157^ for adult kidney tissue DNAse-seq (ENCFF132KNJ), CTCF ChIP-seq (ENCFF498EED), and histone modification ChIP-seq (H3K27Ac - ENCFF584JKX, H3K4me1 - ENCFF955PYL, H3K4Me3 - ENCFF349XHZ) peaks. These files were downloaded from the ENCODE portal under the listed accessions and converted from GRCh38 to GRCh37 coordinates using UCSC liftOver^158^. VEP was also used with adult kidney tissue Cap Analysis of Gene Expression (CAGE) peaks from FANTOM5^159^. These were obtained from the set of ‘Robust human CAGE peaks’, downloaded from the FANTOM5 portal, which were filtered to retain peaks with non-zero read count in kidney tissue. Transcription factor binding sites were obtained from the Ensembl regulatory build using VEP option ‘--regulatory’. VEP was also provided with a BED file of adult kidney tissue chromatin states, produced using an 18-state chromHMM^160^ model by EpiMap^161^. Adult kidney chromatin states were downloaded from the EpiMap portal (accession: BSS01097) and summarized into 6 broader chromatin definitions (enhancer, promoter, transcribed, repressed, heterochromatin, quiescent). Variant scores were also uploaded to VEP for base-wise conservation (phyloP-100way^162^ – calculated across 100 mammalian species) and predicted deleteriousness (CADD^163^ - Combined Annotation Dependent Depletion). PhyloP scores were downloaded from the UCSC genome browser^158^in bigWig format and provided to VEP with ‘--custom’, CADD scores (v1.4) for gnomAD SNPs were downloaded from the CADD website in VCF format and provided to VEP using the CADD plugin.

The enrichment for binary consequence (e.g. missense, synonymous, splice), context, epigenomic and chromatin state annotations, in lead kidney pSNPs (n = 243) was tested using a two-sided Fisher’s exact test and the reference of non-pSNPs (n = 897,745, defined by *P* > gene-specific threshold for all 243 proteins). The *P* values were BH-corrected for the number of comparisons. For CADD and phyloP annotations, lead kidney pSNP and non-pSNP scores were compared using two-sided Mann-Whitney U tests.

The enrichment analysis for gene ontology biological processes in kidney pGenes was performed using clusterProfiler^164^ ‘enrichGO’, against a background of all of 5,764 proteins tested for kidney pQTL. The biological processes with BH-corrected *P* < 0.05 were defined as enriched. For visualization, top enriched pathways with *FDR* < 0.01 were selected and condensed to a set of non-redundant pathways using clusterProfiler ‘simplify’.

### Investigations of shared kidney pQTL and eQTL signals

To investigate the concordance of kidney *cis*-pQTL and *cis*-eQTL effects, we first identified 203 kidney eGenes/pGenes with significant eQTL effects for expression of the transcript, and significant pQTL effects for abundance of the encoded protein. Bayesian colocalization tests were then performed between pQTL and eQTL within their *cis*-windows using the coloc^151^ R package with the default parameters. Colocalization was defined as PP H4 > 0.8. To further investigate the consistency of the signals, we defined ‘eQTL-supported pQTL’ where the lead pSNP exhibited a significant (*FDR* < 0.05) eQTL effect in the same direction as the pQTL effect. The 150 eQTL-supported pQTLs and the 93 remaining pQTLs without eQTL support were then tested for functional enrichment compared to non-pSNPs as previously described.

### Identification of CKD-pGenes

CKD-pGenes were defined as protein partners of molQTL meeting any of the three criteria: (1) colocalization (PP H4 > 0.8) between a pQTL and GWAS trait, (2) colocalization (PP H4 > 0.8) between an eQTL and GWAS trait and a statistically significant directionally consistent pQTL effects, or (3) colocalization (PP H4 > 0.8) between an kidney eQTL and GWAS trait and a significant positive correlation between mRNA of the gene and renal abundance of the protein (*FDR* < 0.05).

### Multi-trait colocalization

As a downstream analysis of selected genes (*KIF12*, *GSTA1*, *OSR1*, *GRB10* and *CA12*), multi-trait colocalization was performed using the HyPrColoc^44^ R package. For each gene, we examined relevant renal *cis*-molQTLs, the CKD-defining traits, and additional European-ancestry GWAS summary statistics (OSR1 – kidney volume^66^, CA12 – serum urate^165^). Harmonized GWAS summary statistics in hg19 build were obtained for kidney volume (N = 32,860) and serum urate (N = 677,373) from GWAS catalog under the accessions GCST90016670 and GCST90319906 respectively., Multi-trait colocalization was assessed using uniform priors and equal regional and alignment thresholds of 0.9 (equivalent to PP H4 ≈ 0.8 in pairwise colocalization) and default priors in HyPrColoc. Colocalization was defined as posterior probability and regional probability both > 0.8 for the resulting trait cluster. Further sensitivity analysis was performed by varying the values for conditional colocalization prior (p_c_ = 0.05, 0.02, 0.01, 0.005) and colocalization threshold (regional threshold = alignment threshold = 0.6, 0.7, 0.8, 0.9, 0.95, 0.99) using the HyPrColoc ‘sensitivity.plot’ function.

### *Cis*-expression Quantitative Trait Methylation analysis

To uncover the identity of kidney *cis*-eQTMs, we tested 369,175 CpG kidney methylation profiles for association with normalized expression of 19,427 nearby genes (TSS < 1Mb) in 366 kidney samples (350 – from HKTR, 16 – from TCGA). Here, we used the full methylation dataset which was generated, quality-controlled, and normalized as described above in the description of *cis*-mQTL analysis. The gene expression profiles were extracted from the normalized mRNA abundance values for corresponding samples from the master kidney gene-expression dataset (645 samples) generated using quality-controlled and normalized data, as described above in the section on *cis*-eQTL analysis.

Our *cis*-eQTM analysis was performed using QTLtools^147^ v1.3, adjusting for the effects for age, sex, study and 3 genetic principal components. *Cis*-eQTM associations were also adjusted for unknown confounders by regressing out the effects of 30 methylation principal components and 80 expression principal components from the kidney DNA methylation and gene expression profiles respectively (using the ‘QTLtools correct’ command). We used the number principal components based on the optimization selection process, as reported above in the respective section of kidney mQTL and eQTL analysis. QTLtools permutation tests, nominal tests, and *P* value corrections were performed using the same parameters and thresholds as in the molQTL analysis.

### Functional annotations of kidney *cis*-mQTL, *cis*-eQTM and kidney CKD-mQTL

To functionally annotate kidney mSNPs, we performed variant annotation and enrichment tests in line with these conducted in functional characterization of pSNPs. Briefly, we tested 73,518 lead kidney mSNPs for the enrichment of variant annotations compared to 3,592,695 non-mSNPs (*FDR* > 0.05 for the same mCpGs), at an *FDR* of < 0.05 to account for multiple testing.

We characterized the genomic context, epigenomic modifications, and chromatin states associated with mCpGs in adult kidney tissue. This analysis utilised CpG positional annotations derived from the HM450 array annotation file^166^, alongside epigenomic and chromatin state annotations derived from DNase-seq, histone ChIP-seq, and 18-state chromHMM analysis of adult kidney tissue^157,161^. The 18 EpiMap chromatin states were summarized to 12 broader classifications of chromatin function. These kidney-specific annotations were derived from the same BED files used in functional annotation of pSNPs and mSNPs. CpG annotations were defined by intersecting CpG positions with peak/chromatin call co-ordinates. To investigate functional enrichment in CpGs, we performed two-sided Fisher’s exact tests for each annotation, and then BH-corrected *P* values. This method was used to assess the functional enrichment in 218 multi-trait CKD-mCpGs, and 856 single-trait CKD-mCpGs, compared to 58,880 non-colocalizing mCpGs (PP H4 > 0.8 with all tested traits.) We also further evaluated kidney emCpGs by comparing the functional enrichment in 3,048 negative emCpGs (effect < 0, *FDR* < 0.05) and 1,792 activating emCpGs (effect > 0, *FDR* < 0.05), compared firstly to 285,239 non-emCpGs (*FDR* > 0.05 for all emGenes), and then to each other.

### Identification and characterization of CKD-mGenes

Given the enrichment of regulatory function in CKD-mCpGs, we sought to link kidney mCpGs to potential target genes. Integrating data from multiple sources, we first linked mCpGs to candidate genes based on: (1) position within a gene body (derived from the HM450 annotation file^166^); (2) distance to gene (derived from calculating CpG-TSS distances using gene annotations from Ensembl 111 GRCh37, and linking each mCpG to all TSS within 1Mb); and significant eQTM effects (*FDR* < 0.05). To reduce the number of genes assigned to mCpGs, and simplify downstream analysis, we introduced a prioritization strategy that assigned each mCpG to (at most) one target gene. For each CpG, candidate genes were scored by: intragenic position (1 point), distance to TSS (1 – (distance / 1Mb)), and kidney eQTM effect (absolute beta coefficient). Finally, the highest scoring gene was selected as the partnered ‘mGene’.

To characterize the effects of CKD-associated methylation, we defined 493 CKD-mGenes (i.e. genes partnered to CKD-mCpG) and performed gene ontology (GO) over-representation analysis. clusterProfiler^164^ was used to test enrichment of GO biological processes in CKD-mGenes, using a background set of 10,666 mGenes linked (by the process described above) to all 59,954 mCpGs tested for colocalization. Enriched GO terms were defined at *FDR* < 0.05, and 12 top enriched pathways (*FDR* < 0.01) were selected for visualization and further analysis. To investigate the contributions of multi-trait vs trait-specific CKD-mGenes to the enrichment results, we tested enrichment of these 12 pathways in CKD-mGenes divided by trait colocalizations in comparison to the set of non-colocalizing kidney mGenes.

We further identified 51 CKD-mGenes nominated by eQTM effects and tested the enrichment of 251 previously identified CKD-eGenes and CKD-mGenes in comparison to 442 CKD-mGenes without an eQTM by two-sided Fisher’s exact test.

### Identification and characterization of developmental kidney CKD-mQTL

Developmental CKD-mGenes’ were defined as 26 CKD-mGenes with known function in renal development listed under GO term ‘GO:0072001 - Renal System Development’ (and accessed using the MSigDB R package^167,168^). These developmental CKD-mGenes were partnered with 68 CKD-mCpGs (termed ‘developmental kidney CKD-mCpGs’). We defined one ‘developmental CKD-mSNP’ for each developmental CKD-mCpG as the most probable colocalizing variant with any CKD-defining trait (i.e. greatest SNP-wise PPH4). Developmental CKD-mSNP’ consisted of 43 unique variants. To test for the enrichment or depletion of eQTL effects in developmental mSNPs, we first identified 42 developmental CKD-mSNPs in our kidney *cis*-eQTL dataset. We then used a two-sided Fisher’s exact test to investigate the prevalence of eQTL effects (*FDR* < 0.05) in comparison to 552 ‘non-developmental CKD-mSNPs’ (which were not linked to developmental CKD-mGenes and were present in our *cis*-eQTL dataset.) The analysis of enrichment for functional annotations in CKD-mCpGs was conducted based on the principles and methods outlined earlier in the analysis of emCpGs. A two-sided Fisher’s exact test was used to test the enrichment of developmental CKD-mCpGs in 218 multi-trait and 856 trait-specific CKD-mCpGs.

### Nomination, prioritization and characterization of CKD kidney genes

To define candidate genes for CKD-defining trait associations we collated the previously defined CKD-eGenes (n = 248), CKD-pGenes (n = 50) and CKD-mGenes (n = 493) into 651 candidate genes prioritized by kidney molecular quantitative trait loci (CKD kidney Genes). Each gene was then scored out of 12 by the number of trait associations (maximum 4) on each molecular level (maximum 3) as defined in previous sections. Briefly, CKD-eGenes were scored 1 or 0 based on eQTL colocalization with the trait (PP > 0.8). CKD-pGenes were scored 1 for pQTL colocalization with the trait; and/or eQTL colocalization with the trait in addition to: (1) directionally consistent eQTL-pQTL effects and/or (2) significant mRNA-protein correlation. Finally, CKD-mGenes (linked to CKD-mCpG by eQTMs and/or positional annotations) were scored 1 if any of their partnered CpGs colocalized with the trait.

We then investigated CKD kidney genes for their novelty as a candidate CKD-GWAS gene, by curating a set of genes previously identified by gene prioritization studies for the 4 CKD-defining traits listed in **Supplementary Table 15**.

CKD kidney genes were also annotated by single gene causes of CKD and renal phenotypes in human and mice. We curated genes with: (1) diagnostic variants found in CKD-patients listed in the supplementary appendix of Groopman et al. 2019^72^ (n = 411); (2), kidney abnormalities reported as a feature of a genetic disease (listed under HPO term: HP:0000077, and accessed from the Human Phenotype Ontology portal^71^, n = 1,451); renal or urinary system abnormalities observed in mice with mutant alleles^70^ (n = 2,024). To obtain the list of mouse phenotype genes, we downloaded mouse-human orthologs with phenotype annotations from the Mouse Genome Informatics^70^ (MGI) portal, using a Human-Mouse Disease Connecter query for the mammalian phenotype ontology ‘MP:0005367’. This identified 2,302 mouse genes associated with renal or urinary abnormalities, which we mapped to 2,024 human orthologs (using the ‘HOM_MouseHumanSequence.rpt’ file downloaded from MGI).

CKD kidney genes were also annotated with drug development information from Open Targets^73^ v25.09.0. We used information obtained from ‘drug/clinical candidates’, ‘drug – mechanism of action’, and ‘drug – indications’ datasets that we downloaded from the Open Targets portal.

### Functional annotation for CKD kidney genes with score > 4

All genes previously categorized as the top 40 CKD kidney genes (score > 4) underwent manual functional annotation in PubMed, Online Mendelian Inheritance in Man (OMIM), and GeneCards to establish gene characteristics, alongside relevance to kidney function or renal disease. Each gene was then linked to a role in the appropriate biological process (metabolism, immunomodulation, intracellular signaling, transport, cell signaling, mRNA editing, cytoskeleton, transcription, detoxification, intracellular transport, transmembrane transport, cell cycle).

### Genetically-regulated expression prediction model and phenome-wide association analysis of renal GRB10 expression

We imputed genetically predicted kidney mRNA expression (GReX) of *GRB10* for 337,350 unrelated individuals of White European ancestry in the UK Biobank^109^ using a pre-calculated kidney expression prediction model^17^. We then performed a GRB10-stratified PheWAS in these individuals 193 quantitative phenotypes derived according to a previously published trait-categorization method^169^. Phenotype processing followed the PEACOK pipeline^169^. Briefly, quantitative traits were rank-based inverse-normal transformed and excluded if N < 500. Associations between imputed *GRB10* kidney expression and quantitative traits were tested by linear regression - all models included age, sex, UK Biobank genotyping array and the top 10 genetic principal components as covariates. Multiple testing was controlled at *FDR* < 0.05.

### Phenome-wide analysis of CA12 genetic variant

We conducted a PheWAS of rs1043256 within *CA12* in the same 337,350 unrelated UKBB individuals of European ancestry. Included in the analysis were 193 quantitative traits/phenotypes identified following the previous trait categorization method^169^. The association analysis between *CA12* variant and each phenotype was carried out using linear regression with age, sex, UK Biobank genotyping array and the top 10 genetic principal components as covariates under additive model of inheritance. Multiple testing was controlled at *FDR* < 0.05. The PheWAS analysis was implemented using the R package PEACOK^169^.

### Conditional analysis and colocalization for CA12

To separate primary and secondary signals in the *CA12* locus, eGFRcys associations were conditioned on rs2729805 (the lead eGFRcys SNP, suspected to mask colocalization with the CA12 eQTL). Approximate conditional analysis was performed using ‘GCTA cojo’^170^, using LD information from European-ancestry UKBiobank participants as a reference. HyPrColoc was then used to assess multi-trait colocalization with the conditioned-eGFRcys associations, and sensitivity analysis was performed by varying the values for conditional colocalization prior (p_c_ = 0.05, 0.02, 0.01, 0.005) and colocalization threshold (regional threshold = alignment threshold = 0.6, 0.7, 0.8, 0.9, 0.95, 0.99) using the HyPrColoc ‘sensitivity.plot’ function.

### *In silico* prediction of CA12 loss-of-variant effects

Functional prediction analyses were performed using an integrative approach. First, all isoforms were analysed with InterPro^87^ in order to identify functional domains and signatures. Presence/absence of transmembrane regions were confirmed with DeepTMHMM^89^. Finally, Quick2D^88^, NetSurfP-3.0^90^ and CAID^91^ were used to validate that the secondary structure and solvent accessibility of the isoforms were compatible with the functional domain predictions. A model structure for the O43570-1 isoform was downloaded from the AlphaFold Protein Structure Database^171^ (https://alphafold.ebi.ac.uk/)(AF-O43570-F1-model_v4). ColabFold^172^ was used to build an AlphaFold2^92^ model structure for the equivalent novel isoform. Both model structures were analysed and optimised with MolProbity^173,174^ by adding hydrogens and correcting bad flips. ChimeraX^175^ was used to superimpose both optimised model structures and to generate figures coloured based on DeepTMHMM predictions, Coulombic electrostatic potentials, hydrophobicity^174,176^ and B-factor.

### *In vitro* validation of CA12 loss-of-function variant

*In vitro* analyses used HK-2 cells (Caltag Medsystems, T0011004), a proximal tubule cell line derived from an adult male kidney^177^. Cells were maintained in K-SFM supplemented with bovine pituitary extract and EGF (Invitrogen, 17005-042) at 37°C with 5% CO_2._

Plasmids for HiBiT reporter assays were designed to express the expected coding sequences of either rs148438059-C (with exon 10) or rs148438059-T (without exon 10) with a C-terminal HiBiT tag fusion (**Supplementary Fig. 4**). Tagged coding sequences were synthesised as custom gBlock sequences (IDT) then inserted into the pcDNA3.1 expression plasmid backbone by HiFi DNA assembly (NEB, E5520S). Plasmid identities were confirmed by Sanger sequencing and DNA was purified using SmartPure plasmid (Eurogentec, SK-PLPU-100) or NucleoBond Xtra Midi (Macherey-Nagel, 740410.50) kit.

HK-2 cells (9×10^5^/reaction) were nucleofected with a mix of HiBiT reporter plasmid (2.5μg), a CMV-luciferase plasmid (500ng) and a constitutive GFP plasmid (300ng) in buffer SF using a 4D Nucleofector (Lonza, programme EH100). Cells were seeded into 6-well tissue culture plates to recover for two days, then nucleofection efficiency was confirmed by visualisation of GFP fluorescence and cells were detached using trypsin-EDTA (Sigma-Aldrich, T3924) for reseeding into 5-6 replicate wells of a white 96-well plate. Cells were allowed to recover for one day prior to luciferase assay. Measurements were collected for each well using a Tecan Spark platereader after (1) addition of luciferin (Promega, E160B) to detect firefly luciferase, (2) addition of NanoGlo HiBiT extracellular reagent (Promega, N2420) then (3) addition of Triton X-100 to a final concentration of 0.1% to lyse cells. HiBiT measurements were restricted to a spectral window of 360-485nm to prevent residual firefly luciferase detection and signal identity was confirmed by spectral deconvolution using the luminescence scan acquisition mode.

### Analysis of CA12 loss-of-function variant phenotype associations

We investigated associations between the rs148438059-T allele and in European ancestry participants from UK Biobank^109^ (UKBB), AllOfUs^93^ (AOU), Million Veterans Program^94^ (MVP) and CKDGen^95^ for selected metabolic (sodium, potassium and chloride in blood and urine) and renal phenotypes (eGFRcr, eGFRcys, BUN, urate, CKD, gout). For UKBB and AoU, we extracted genotype dosages for the rare CA12 loss-of-function allele (rs148438059-T) in individuals of European ancestry (UKB = 408,567, AOU = 232,404) together with the corresponding phenotypes. These included serum biochemistry (sodium, potassium, chloride, urate, creatinine, cystatin C and urea), alongside urine analyses (sodium and potassium) as well as disease diagnoses from ICD codes (CKD and gout). CKD and gout were defined using first occurrence summary-level diagnoses in UKB (ICD-10 codes: N18 and M10, respectively) and in AOU, diagnoses were retrieved for all participants with matching concept identifiers. Measures of eGFR were derived from creatinine (eGFRcr) and cystatin C (eGFRcys) using the race-free CKD-EPI equations^105^, and blood urea nitrogen was derived from serum urea measurements. Quantitative traits were normalised using rank-based inverse-normal transformation and disease diagnoses were re-coded as binary phenotypes.

To examine associations between rs148438059-T dosage and the phenotypes in UKBB, we applied dominant linear and logistic regression models to quantitative and binary phenotypes^178^. All models were adjusted for age, sex and the first 10 genetic principal components with genotype array applied as a UKB study-specific covariate. Firth’s correction was applied to logistic models to account for case-control imbalance where appropriate using the brglm2 R package^179^. For MVP, phenotype associations (eGFRcr, BUN, serum sodium, serum potassium, serum chloride, CKD and gout) for rs148438059-T were extracted from summary statistics generated by Verma *et al.* (2024)^94^ and downloaded through the dbGAP portal^180^. For CKDGen, eGFRcr and eGFRcys associations for rs148438059-T were extracted from summary statistics generated by Li *et al.* (2017)^181^ and downloaded from the CKDGen portal^95^.

A fixed-effect inverse-variance weighted meta-analysis was conducted to combine effect estimates for each phenotype in all cohorts where measurements were available using METAL software^112^. Cochran‘s Q tests in METAL found no evidence of heterogeneity in any of the analyses (P > 0.05). The level of statistical significance for meta-analysed associations was defined by the Bonferroni threshold *P* < 0.0045.

## Data Availability

GWAS associations for lead variants in each locus region for eGFRcr, eGFRcys, BUN and UACR are provided in Supplementary tables 1-4. Lead molQTL associations are provided in Supplementary tables 10-13 for mQTL, eQTL, pQTL and mirQTL. Additional data are available from the authors upon reasonable request.

## Code Availability

All analyses make use of preexisting code and software packages, which are fully described and referenced in the Methods section.

## Acknowledgements

This work was supported by British Heart Foundation grants (PG/19/16/34270, PG/22/10957 and PG/24/11687), to M.T., British Heart Foundation Research Excellence Award (RE/24/130017), National Institute for Health and Care Research (NIHR) Manchester Biomedical Research Centre (BRC) (NIHR203308), (to M.T., A.P.M. and B.K.). A.E. was supported through National Institute for Health and Care Research (NIHR) Manchester Biomedical Research Centre (BRC) (NIHR203308) and received pump priming funding from the British Heart Foundation Manchester Research Excellence Award (RE/24/130017) and the Manchester BRC (NIHR203308). A.E. received a travel grant from Kidneys for Life. Y.S is supported by China Scholarship Council Innovation Platform with SJTUSM (Project ID: 202306230019). A.C.L. is supported by an Intermediate Fellowship from Kidney Research UK (INT_002_20220705) and received pump priming funding from British Heart Foundation Manchester Research Excellence Award (RE/24/130017), Manchester BRC (NIHR203308) and University of Manchester ‘Healthier Futures’ Platform.

This project was supported by UK Biobank application (46114). The views expressed are those of the author(s) and not necessarily those of the NIHR or the Department of Health and Social Care.

Parts of this work were presented at the annual meeting of the American Society for Human Genetics 2024 and the European Renal Association congress 2025 (European Renal Association 2025 outstanding abstract submission to A.E.). We thank the University of Manchester Genome Editing Unit, especially Dr Alis Hales, for technical expertise.

We dedicate this work to Professor Ewa Zukowska-Szczechowska – professor of nephrology, an outstanding mentor to the next generation of scientists and clinicians and a tireless champion for excellence in kidney research.

## Ethics Declaration

The studies adhered to the Declaration of Helsinki and were approved/ratified by the Bioethics Committee of the Medical University of Silesia (Katowice, Poland), Bioethics Committee of Karol Marcinkowski Medical University (Poznan, Poland), Ethics Committee of University of Leicester (Leicester, UK), University of Manchester Research Ethics Committee (Manchester, UK) and National Research Ethics Service Committee Northwest (Manchester, UK). Informed written consents were obtained from all individuals recruited (for the deceased donors, the consent was obtained in line with the local governance, e.g. from the family members).

---

**Extended Data Fig. 1.**
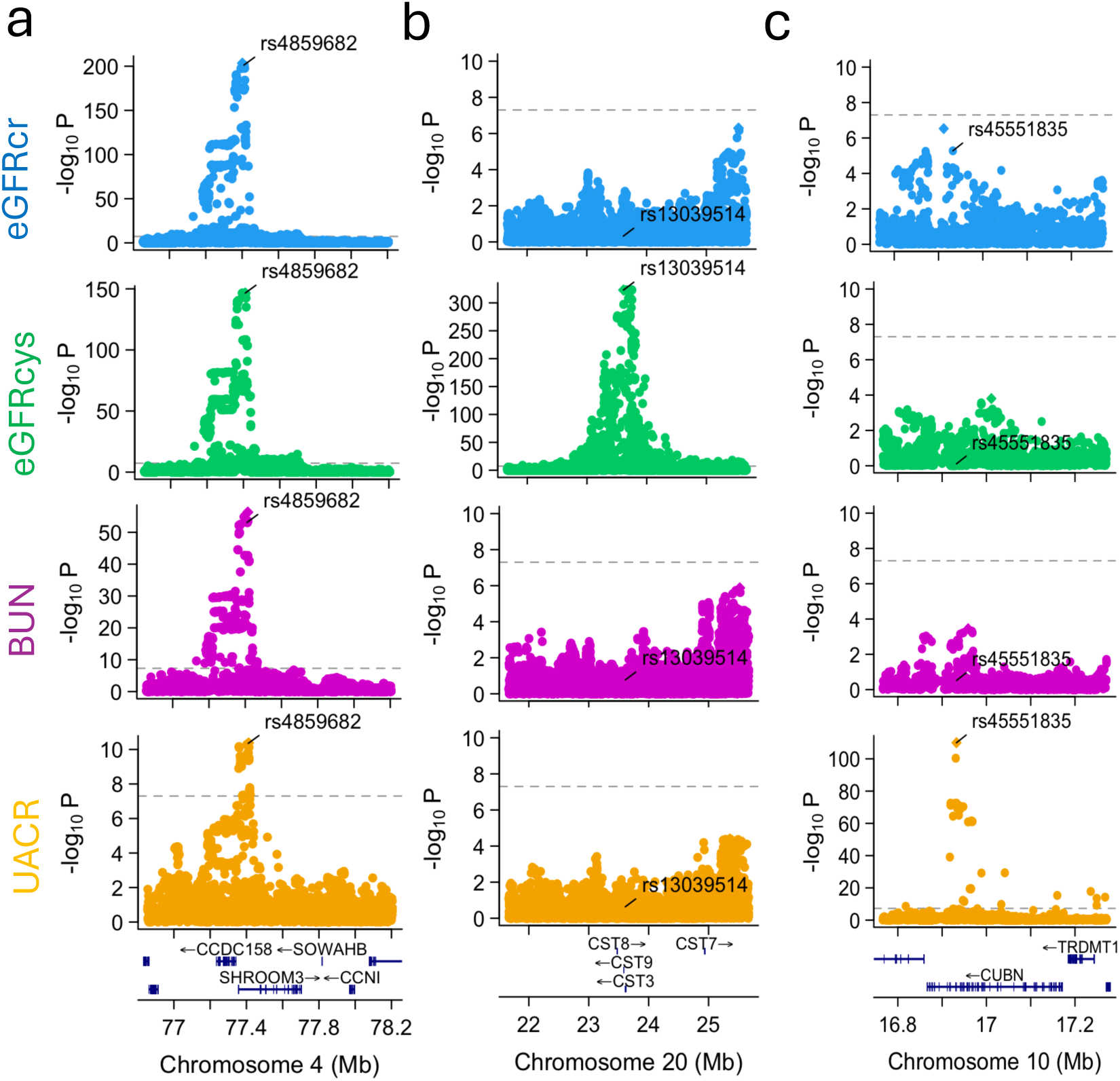
Examples of shared and trait-specific loci. Each panel shows LocusZoom plots for the 4 CKD-defining traits, where the *x* axis shows chromosomal position, and the *y* axis shows −log_10_P values. Loci are annotated with the variant which has the smallest *P* value across all four GWAS. **A** shows the multi-trait *SHROOM3* locus, **B** shows the eGFRcy s-specific cystatin gene cluster locus, and **C** shows the UACR-specific *CUBN* locus.

**Extended Data Fig. 2.**
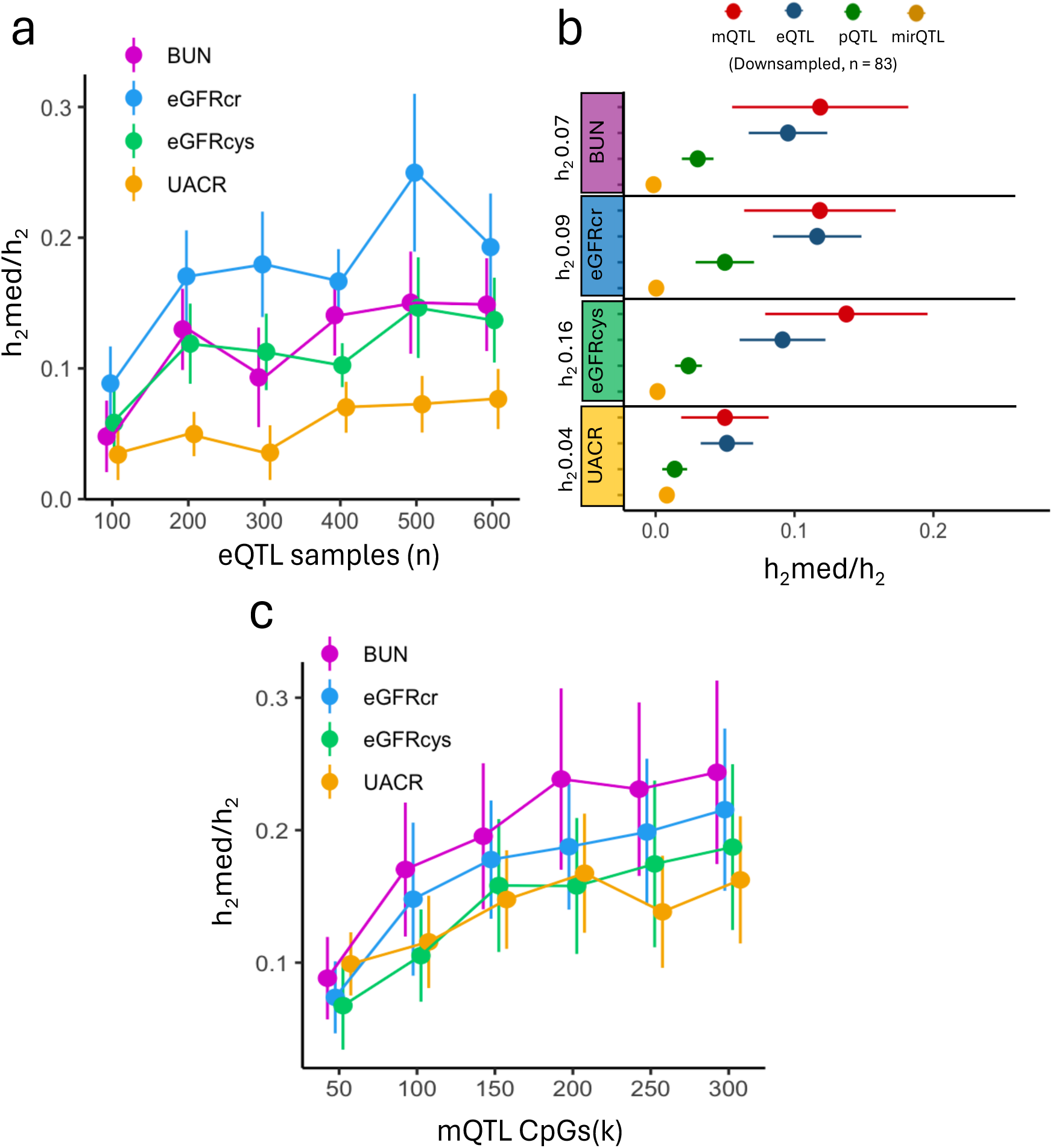
MESC sensitivity analyses. **A.** Estimates of mediated heritability (h_2_med/h_2_) calculated using down-sampled eQTL datasets for 4 CKD-defining traits by MESC. *Y* axis shows estimates of h_2_med/h_2_ for each eQTL sample size (*x* axis). Points are coloured by CKD-defining trait (blue – eGFRcr, green – eGFRcys, purple – BUN, yellow – UACR). **B.** Estimates of h_2_med/h_2_ by molQTL, downsampled to 83 random samples in each dataset. *X* axis shows h_2_med/h_2_ estimates and standard errors for each CKD-defining trait (*y* axis). Points are coloured by molQTL datatype (red – mQTL, green – eQTL, blue – pQTL, yellow – mirQTL). **C.** Estimates of h_2_med/h_2_ by mQTL (n = 366) for varying numbers of CpGs. *Y* axis shows estimates of h_2_med/h_2_ for each mQTL CpG size (*x* axis). Points are coloured by CKD-defining trait (blue – eGFRcr, green – eGFRcys, purple – BUN, yellow – UACR).

**Extended Data Fig. 3.**
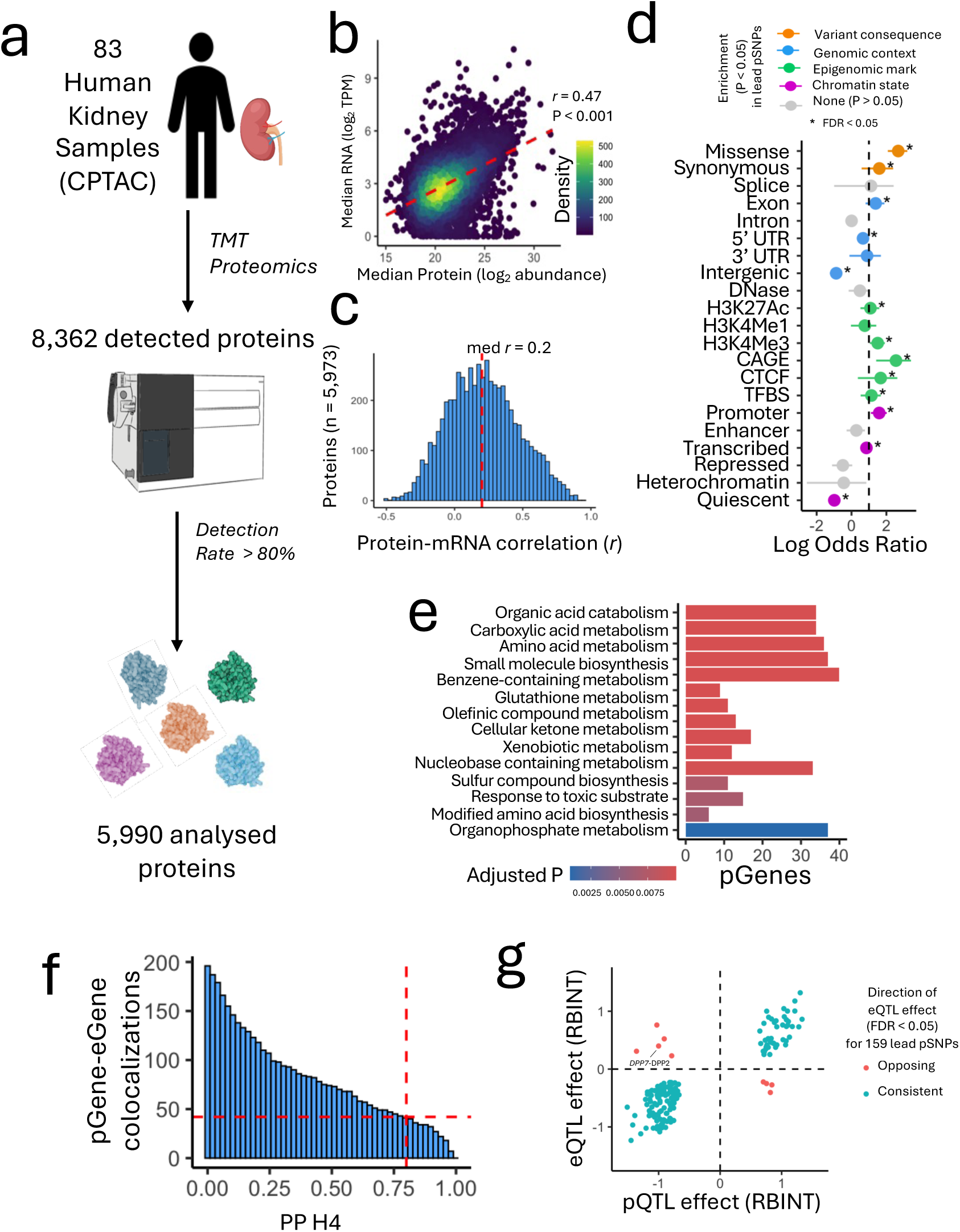
Integration of 83 genomes, transcriptomes and proteomes in CPTAC kidney samples. **A.** Schematic of proteomics analysis. Kidney samples were examined by tandem mass-spectrometry (TMT). Proteins were filtered for >80% detection in the 83 samples, yielding 5,990 proteins for downstream analysis. **B.** Correlation between median protein abundance and median mRNA abundance calculated across 76 CPTAC samples with matched proteomics and RNA-seq transcriptomic data. *X* axis shows log_2_ normalized median protein abundance and *y* axis shows median log_2_ TPM normalized mRNA abundance. Points, representing 5,973 proteins, are coloured by density of values. **C.** Individual correlations between log_2_ normalized protein abundance and log_2_ normalized mRNA abundance for the 5,973 proteins in 76 samples. Histogram shows distribution of Pearson correlation coefficients (*x* axis). **D.** Enrichment of functional annotations in 243 lead pSNPs, compared to 897,745 non-pSNPs. *X* axis shows enrichment log odds ratio for each functional annotation (*y* axis) calculated by two-sided Fisher’s exact test. Points are coloured by annotation category, unless there was no significant enrichment/depletion (P > 0.05). ‘*’ indicates significance after correction for multiple testing (*FDR* < 0.05). UTR – untranslated region, CAGE – cap analysis of gene expression, TFBS – transcription factor binding site. **E.** Top enriched non-redundant GO pathways (*FDR* < 0.01) in 243 pGenes compared to 5,521 non-pGenes. **F.** Number of proteins with colocalized pQTL and eQTL signals (*y* axis) when altering the threshold for posterior probability of colocalization (PP H4, *x* axis) for 203 proteins with both eQTL and pQTL. Dashed line shows number of colocalizations (n = 43) for the default 0.8 threshold. **G.** Relationship between kidney eQTL and pQTL coefficients for 159 lead pSNPs with significant eQTL effects. Points are coloured by consistent (blue) or opposing (red) directionality. eQTL and pQTL effect sizes are rank-based inverse normal transformed (RBINT) changes in normalized mRNA and protein abundance respectively.

**Extended Data Fig. 4.**
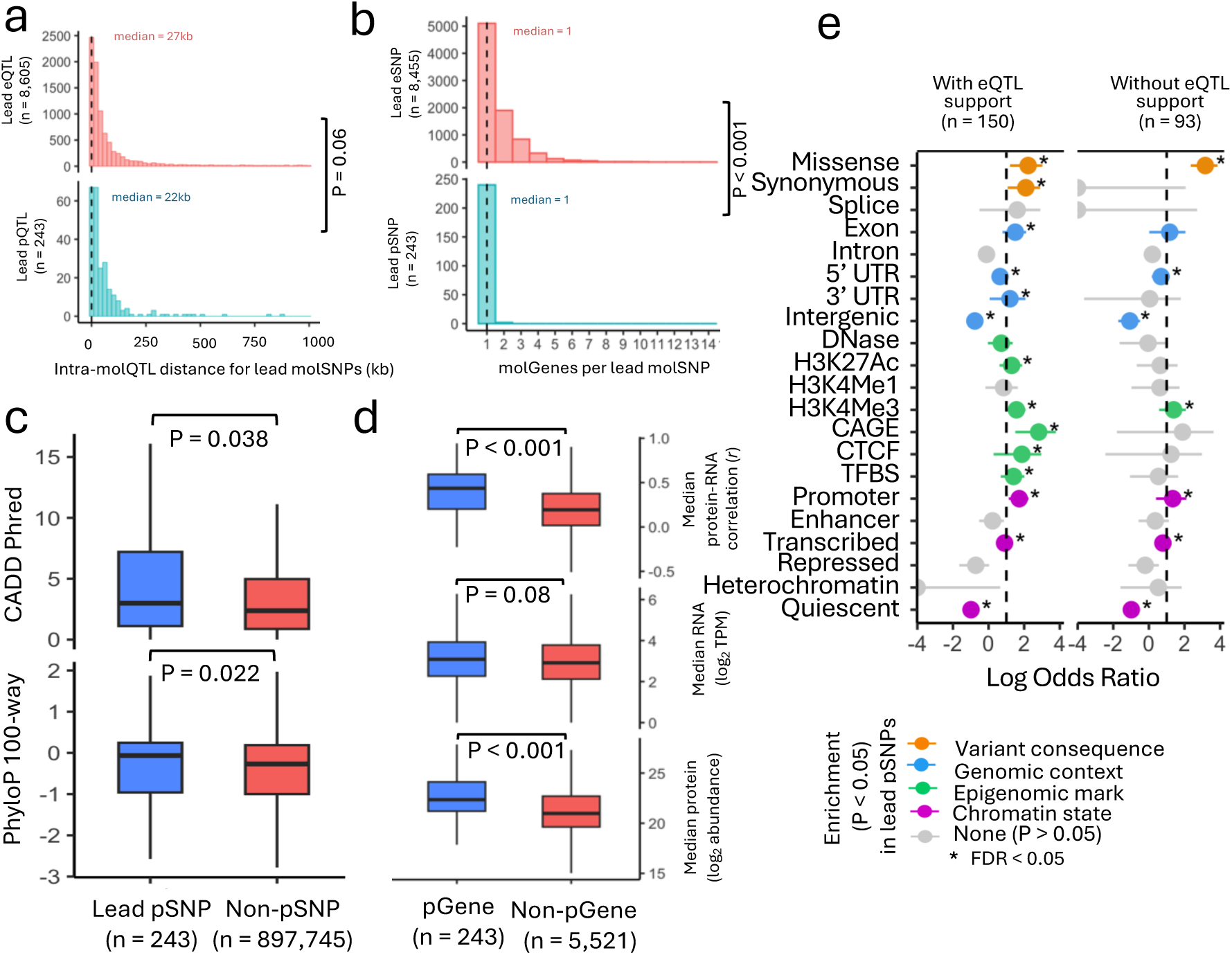
Functional characterization of kidney pQTL. **A.** Distributions of intra-molQTL distances (*x* axis) between lead kidney eSNPs and eGenes (top) and lead kidney pSNPs and pGenes (bottom), *P* value from two-sided Mann-Whitney U test. **B.** Distributions of eGene/pGene partners (*x* axis) for lead pSNPs (top) and lead pGenes (bottom), *P* value from two-sided Mann-Whitney U test. **C.** Boxplots showing distributions of scores for predicted SNP deleteriousness (CADD scores - top), and SNP conservation (phyloP 100-way - bottom) for lead kidney pSNPs compared to non-pSNPs, *P* value from two-sided Mann-Whitney U test. **D.** Boxplots showing distributions of median mRNA-protein correlations (above), median normalized mRNA abundance (middle), and median normalized protein abundance (bottom) for kidney pGenes vs non-pGenes, *P* value from two-sided Mann-Whitney U test. **E.** Enrichment of functional annotations in lead kidney pSNPs with eQTL support (i.e. with directionally consistent eQTL, left), and lead kidney pSNPs without eQTL support (i.e. with no significant eQTL effect or directionally inconsistent eQTL effect, right). Enrichment calculated by two-sided Fisher’s exact test compared to 897,745 non-pSNPs. *X* axis shows enrichment log odds ratio for each functional annotation, points are coloured by annotation category, unless there was no significant enrichment/depletion (P > 0.05). ‘*’ indicates significance after correction for multiple testing (*FDR* < 0.05). UTR – untranslated region, CAGE – cap analysis of gene expression, TFBS – transcription factor binding site).

**Extended Data Fig. 5.**
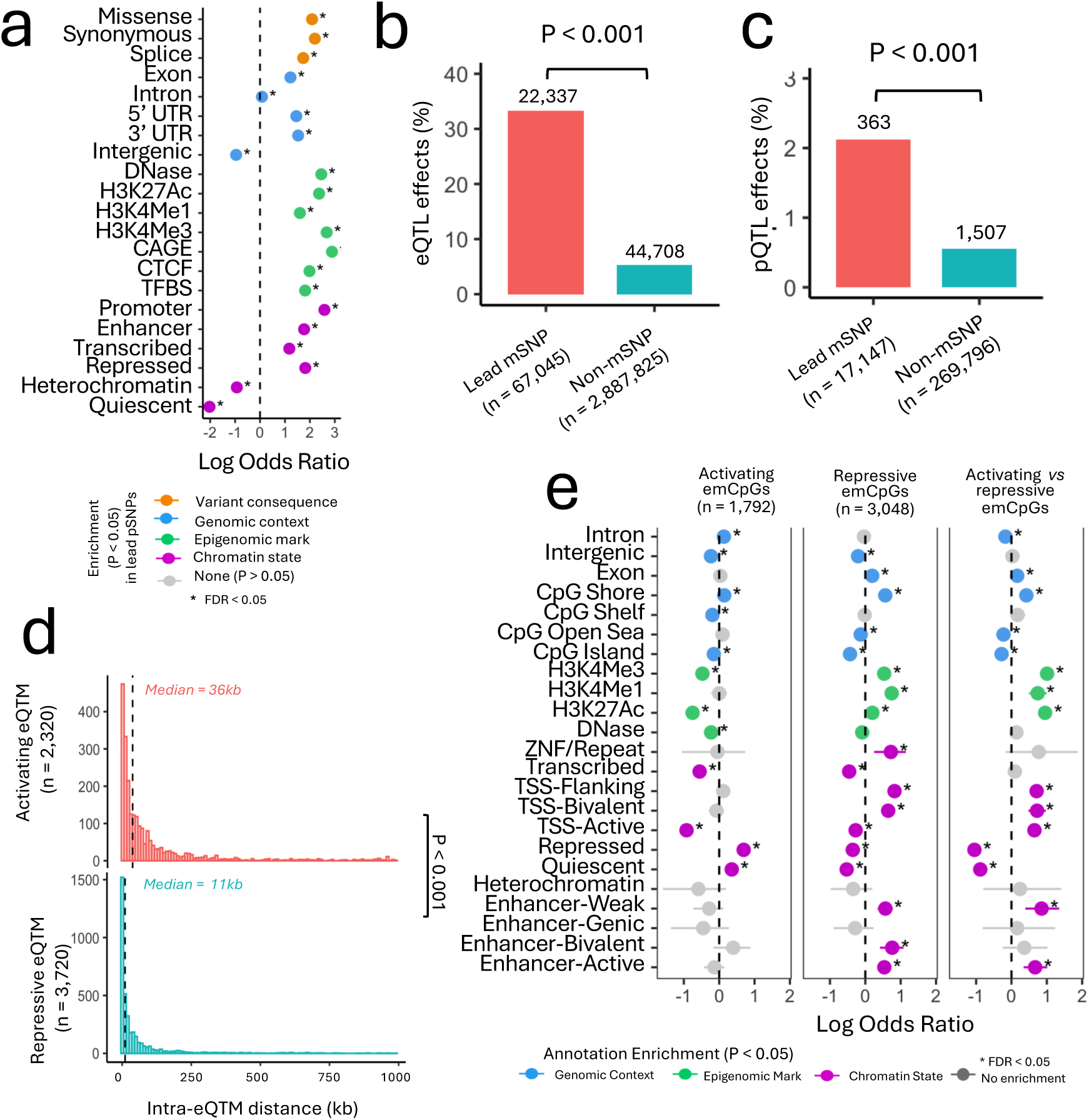
Functional characterization of kidney mQTLs and eQTMs. **A.** Enrichment of functional annotations in 73,518 lead kidney mSNPs, compared to 3,592,695 non-mSNPs. *X* axis shows the log odds ratio (where log odds ratio > 1 indicates enrichment and < 1 indicates depletion) for each functional annotation (*y* axis). Points are coloured by annotation category, unless there was no significant enrichment/depletion (P > 0.05), ‘*’ indicates significance after correction for multiple testing (*FDR* < 0.05) calculated by two-sided Fisher’s exact test. UTR – untranslated region, CAGE – cap analysis of gene expression, TFBS – transcription factor binding site**. B.** Percentage of variants with significant kidney eQTL effects (*FDR* < 0.05, *y* axis) for lead kidney mSNPs and non-mSNPs tested by eQTL analysis (*x* axis). *P* value from two-sided Mann-Whitney U test. **C.** Percentage of variants with significant kidney pQTL effects (at *FDR* < 0.05 and *P* < gene-specific threshold, *y* axis) for lead kidney mSNPs and non-mSNPs tested by kidney pQTL analysis (*x* axis). *P* value from two-sided Mann-Whitney U test. **D.** Distributions for intra-eQTM distances (*x* axis) for 2,320 activating eQTM (top) and 3,720 repressive eQTM (bottom). *P* value from two-sided Mann-Whitney U test, dashed lines show median distances. **E.** Enrichment of CpG annotations in activating and repressive emCpGs. There are fewer emCpGs than eQTM since each emCpG may be partnered to multiple genes (range 1-4). *X* axis shows the log odds ratio of enrichment for each annotation (*y* axis), where log odds ratio > 0 indicates enrichment and < 0 indicates depletion. Points are coloured by annotation category (if *P* < 0.05), and lines show 95% confidence intervals. Left panel shows enrichment calculated in 1,792 activating emCpGs compared to 285,239 non-emCpGs (with *P* > gene-specific threshold for all emGenes), middle shows enrichment in 3,048 repressive emCpGs compared to non-emCpGs, and right shows enrichment in uniquely repressive emCpGs (compared directly to activating emCpGs. These analyses excluded 217 emCpGs with both activating and repressive effects. *P* values calculated by a two-sided Fisher’s exact test, ‘*’ indicates significant enrichment after correction for multiple testing (*FDR* < 0.05).

**Extended Data Fig. 6.**
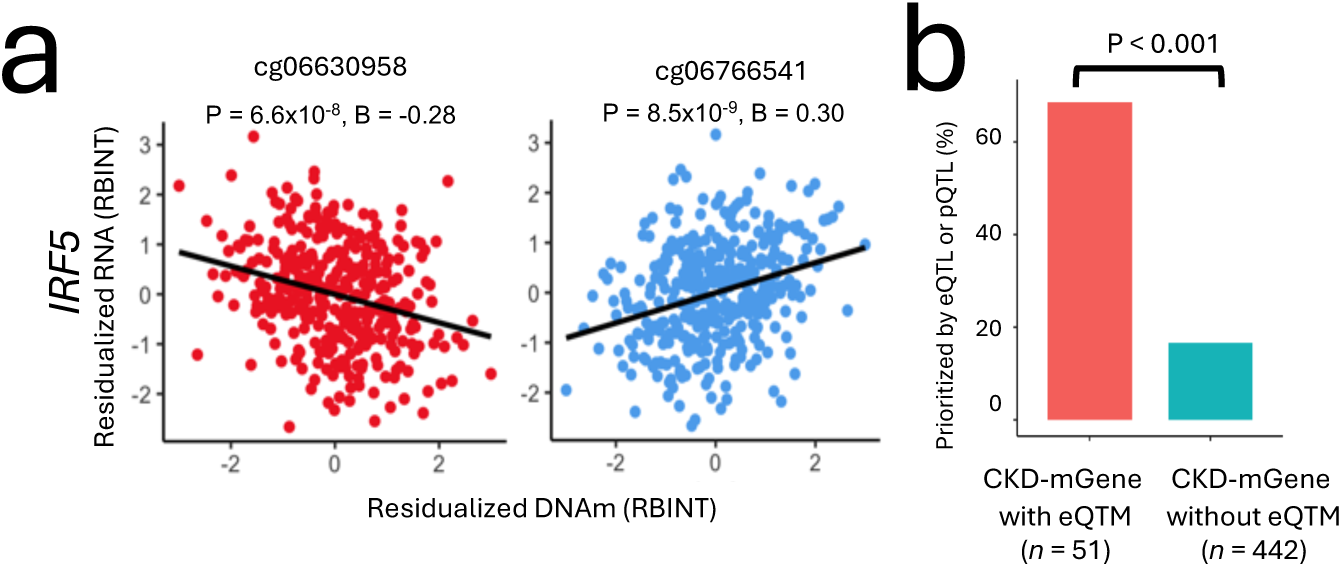
Examples of CKD-eQTMs and enrichment for CKD kidney eGenes and pGenes. **A.** Negative (left) and positive (right) eQTM for the CKD-emGene *IRF5*. *X* axis shows *IRF5* mRNA expression (normalized, rank-based inverse normal transformed (RBINT) and corrected for 80 transcriptomic principal components) and *y* axis shows CpG methylation (normalized, *RBINT* and corrected for 20 DNA methylation principal components) in each of the 366 samples with matched renal transcriptomic and DNA-methylation information (points). Effect size and *P* values from eQTM analysis. **B.** Enrichment of genes prioritized by kidney eQTL or kidney pQTL (union of CKD-eGenes and CKD-pGenes, *N* = 251) in CKD-emGenes. *X* axis shows proportion of CKD-mGenes that are also CKD-eGenes/CKD-pGenes for CKD-mGenes with eQTMs (i.e. CKD-emCpGs, *N* = 51) and CKD-mGenes without eQTMs (n = 442, *y* axis). *P* value calculated by two-sided Fisher’s exact test.

**Extended Data Fig. 7.**
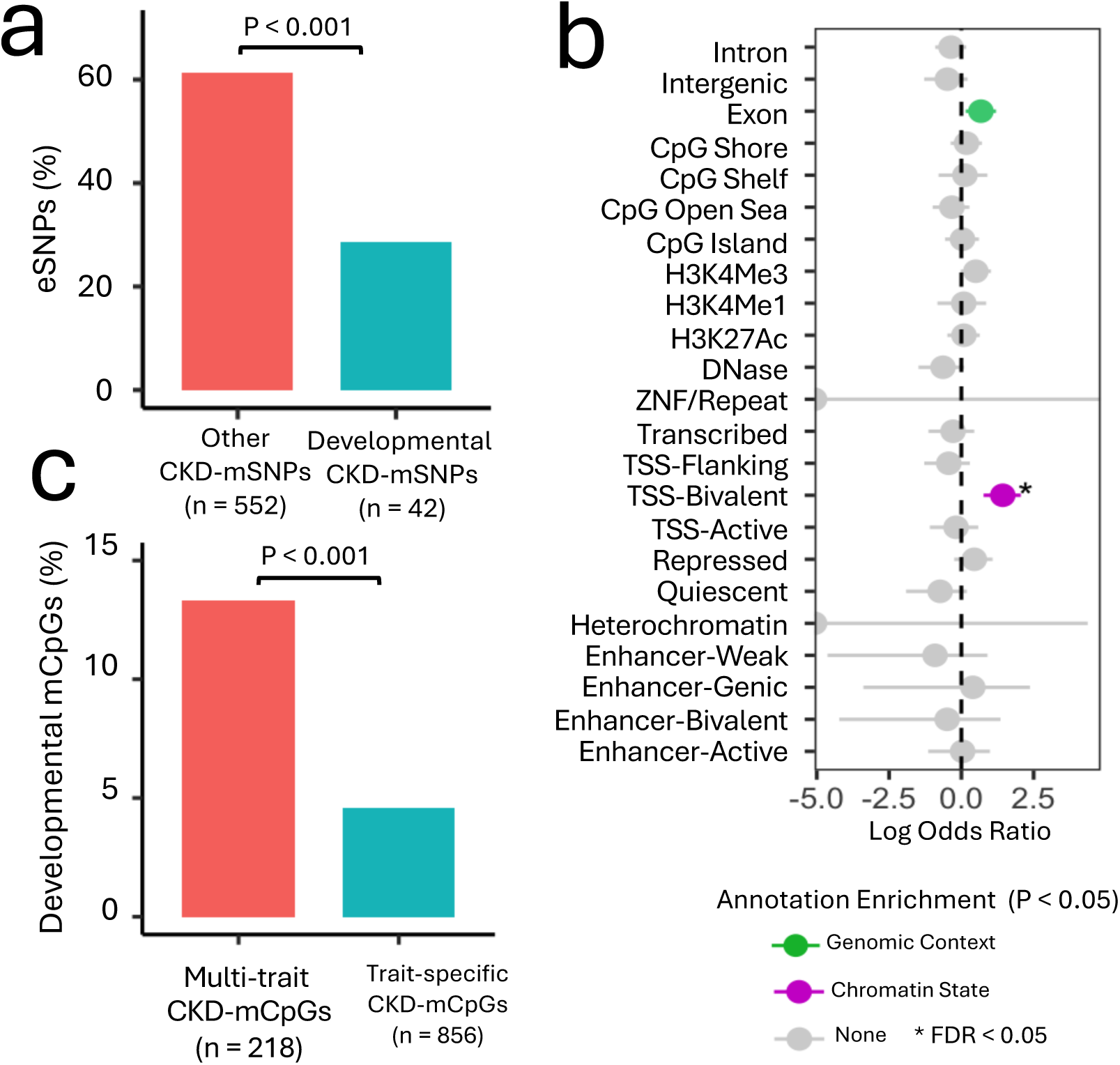
Characterization of developmental CKD-mQTL. **A.** Depletion of eSNPs from developmental CKD-mSNPs. Y axis shows the percentage of CKD-mSNPs with eQTL effects (*FDR* < 0.05) for 42 developmental CKD-mSNPs and 552 CKD-mSNPs not linked to renal development genes. *P value* from two-sided Fisher’s Exact Test**. B.** Enrichment of CpG annotations in developmental CKD-mCpGs (n = 68) compared to all other CKD-mCpGs not linked to renal development genes (n=1,006). *P* values calculated by a two-sided Fisher’s exact test. *X* axis shows log odds ratio (OR) for each annotation (*y* axis), where a log odds ratio > 1 indicates enrichment, and <1 indicates depletion. Points are coloured by annotation category, and lines show 95% confidence intervals. ‘*’ indicates significant enrichment after correction for multiple testing (*FDR* < 0.05). **C.** Enrichment for developmental CKD-mCpGs in multi-trait CKD-mCpGs. Y axis shows the percentage of developmental CKD-mCpGs for 218 multi-trait CKD-mCpGs and 856 trait-specific CKD-mCpGs. *P* value from two-sided Fisher’s Exact Test.

**Extended Data Fig. 8.**
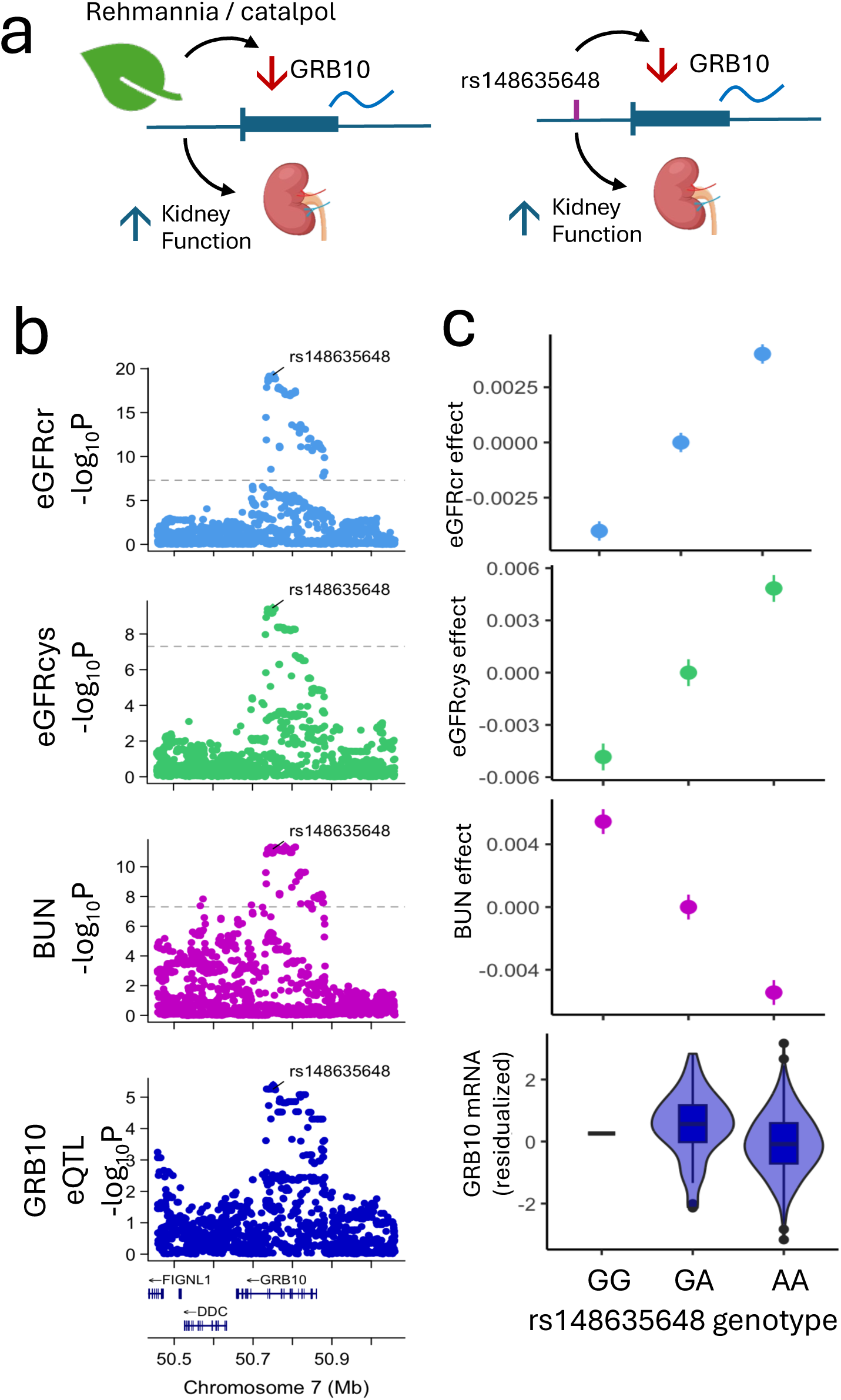
Shared genetic associations between renal GRB10 mRNA, eGFRcr, eGFRcys and BUN. **A.** Schematic showing downregulation of GRB10 expression and improvement of kidney function observed upon treatment with catalpol (active ingredient in the Chinese herbal medicine Rehmannia) in animal studies; consistent with the direction of the shared causal genetic association between reduced renal *GRB10* mRNA and increased kidney function. **B.** LocusZoom plots for eGFRcr, eGFRcys, BUN, and the kidney *cis*-eQTL for GRB10. *X* axis shows chromosomal position, and *y* axis shows −log_10_P values. Loci are annotated with the most likely causal variant. **C.** Effect of rs148635648 genotype on eGFRcr, eGFRcys, BUN and kidney expression of GRB10. For eGFRcr, eGFRcys and BUN, the *y* axis gives the estimated effect of each additional effect allele on the residualized GWAS phenotype. Points show effect size estimates and lines show standard errors. For the GRB10 eQTL, the *y* axis shows the residualized DNA methylation phenotype, and the boxplots show the distribution of values for each genotype.

